# Operational complexity predicts selective non-dissemination within pharmaceutical sponsor portfolios: a retrospective cohort study

**DOI:** 10.64898/2026.05.05.26352331

**Authors:** Ahmed M. Sayed, Phan Thieu Huan, Thuan K. Nguyen, Eman Fathy, Toka Aziz, Duong Van Tho, Nguyen Tien Huy

**Author notes:** Co-corresponding authors: Dr. Nguyen Tien Huy, and Ahmed M. Sayed.

## Abstract

**Background:** Incomplete dissemination of clinical trial results remains an important challenge for research transparency and evidence synthesis. Although prior studies have quantified the overall extent of non-dissemination, less is known about whether trial characteristics observable at registration are associated with subsequent dissemination within sponsor portfolios.

**Methods and findings:** We conducted a retrospective cohort study of 17,537 completed interventional clinical trials registered on ClinicalTrials.gov between 2007 and 2024 across the 20 largest global pharmaceutical companies. We developed the Operational Complexity Index (OCI), a composite measure derived from planned enrollment, facility count, and geographic scope, and examined its association with trial dissemination using multivariable logistic regression and time-to-event analyses.

Higher OCI was associated with greater odds of dissemination (adjusted odds ratio [aOR] = 2.40, 95% CI 2.23–2.60; p < 0.001), with dissemination increasing from 47% in the lowest OCI decile to 95% in the highest. Higher operational complexity was also associated with earlier dissemination; over a 1,095-day horizon, high-OCI trials were disseminated a mean of 310.88 days earlier than low-OCI trials (RMST difference, 310.88 days; 95% CI 300.59–320.96; p < 0.001). This pattern was observed across sponsors, clinical phases, and therapeutic areas. In predictive analyses using registration-time variables, the structural model achieved a cross- validated AUC of 0.816 and a holdout AUC of 0.814, whereas the full model, including sponsor identity, achieved a cross-validated AUC of 0.858 and a holdout AUC of 0.857.

Using benchmark phase-based costing assumptions, the 5,019 non-disseminated trials corresponded to an estimated US$10.94–15.26 billion in sunk research investment.

**Conclusions:** Among trials conducted by the 20 largest pharmaceutical sponsors, greater operational complexity at registration was associated with a higher likelihood of dissemination and earlier dissemination. These findings suggest that aggregate sponsor-level transparency metrics may mask important heterogeneity within sponsor portfolios. Future work should assess whether registration-time trial characteristics can help identify trial subgroups at higher risk of non-dissemination.

**AUTHOR SUMMARY:** *Why was this study done?:*

- Incomplete dissemination of clinical trial results reduces the completeness of the medical evidence base and the public value of research participation.
- Previous studies have described overall rates of trial non-dissemination, but less is known about whether dissemination varies systematically across different types of trials within sponsor portfolios.
- We examined whether trial characteristics available at registration were associated with later dissemination of results among large pharmaceutical sponsors.

*What did the researchers do and find?:*

- We analyzed 17,537 completed interventional clinical trials sponsored by the 20 largest pharmaceutical companies and registered on ClinicalTrials.gov between 2007 and 2024.
- We developed an Operational Complexity Index (OCI) based on planned enrollment, number of facilities, and geographic scope to measure trial operational scale at registration.
- Higher OCI was associated with a greater likelihood of dissemination and earlier dissemination. Dissemination ranged from 47% in the lowest OCI decile to 95% in the highest.
- This pattern was observed across sponsor portfolios, clinical phases, and therapeutic areas, with an average within-sponsor dissemination gap of 40 percentage points between lower- and higher-complexity trials.
- In manual validation of 344 sampled trials, the automated dissemination-classification pipeline achieved 92.1% accuracy.
- Using benchmark phase-based costing assumptions, the 5,019 non-disseminated trials corresponded to an estimated US$10.9–15.3 billion in sunk research investment.

*What do these findings mean?:*

- Dissemination was not uniform across trial types within sponsor portfolios; trials with lower operational complexity were less likely to be disseminated than trials with higher operational complexity.
- Aggregate sponsor-level transparency measures may therefore miss important differences within portfolios.
- Registration-time trial characteristics showed predictive signal for non-dissemination, but whether such information could support monitoring strategies would require prospective validation.
- More complete dissemination of trial results would strengthen the scientific record and improve the public value of clinical research.

## Introduction

Participants in clinical trials help in building knowledge intended to improve care for future patients. This assumption is enshrined in the Declaration of Helsinki(1) and enforced by the ICMJE through the requirement that trial sponsors publish their trial results in a timely manner(2). Failure to disseminate trial results can compromise the completeness of the medical evidence base and reduce the scientific value derived from participant contributions. Non-dissemination remains a substantial problem. Prior studies suggest that nearly one-third of clinical trials funded by drug companies do not report their results once completed(3). In total, it is estimated that over $100 billion is invested annually into research that is never reported(4). Legislative and regulatory efforts have sought to improve trial registration and reporting, although incomplete dissemination persists. For example, the Food and Drug Administration Amendments Act (FDAAA 2007) was enacted in 2007, requiring prospective registration and reporting of results for all U.S.-based trials(5, 6). Following this enactment, the FDA made two additional changes to its regulations regarding the disclosure of clinical trial results in 2016 and 2018(7). However, despite these regulatory improvements, clinical trials continue to go unreported at extremely high rates – even for some of the most well-resourced pharmaceutical companies – indicating that the issue is not a matter of regulatory structure or design, but rather a function of the structural incentives inherent in how trials are designed and executed.

Prior studies have evaluated clinical trials’ silence as a uniform and aggregate phenomenon. These studies have typically measured the percent of a sponsor’s trials that reported results, without evaluating which specific trials were being selectively suppressed and why(8). This approach has resulted in compliance measures that are systematically misleading(9). For example, a company may receive high marks in terms of aggregate transparency ratings while consistently suppressing the results of its smaller, less visible trials. Therefore, researchers are unable to identify which trials are inherently at greater risk of being suppressed due to structural factors. Consequently, regulators are forced to respond in a purely retrospective manner and reactively to clinical trials’ silence, which also prevents regulators from accounting for heterogeneity among clinical trials conducted by a single sponsor.

To address this gap, we have developed the Operational Complexity Index (OCI), a framework that quantifies trial operation complexity at registration. The OCI is intended to quantify the operational footprint of a clinical trial using registration-time characteristics. Specifically, the OCI captures the planned enrollment size of a trial, the number of participating facilities and locations, and the geographic scope of a trial. Although prior work has tethered trial size to dissemination, the OCI extends beyond enrollment size alone to evaluate the degree of geographic dispersion and institutional breadth(10, 11). These dimensions may capture features of trial visibility and operational scale beyond participant volume alone. In model comparisons, an OCI-based structural model showed better predictive discrimination than an enrollment-only model, suggesting that the composite measure captures information beyond enrollment alone.

We hypothesized that operational complexity would be linked to the likelihood of trial dissemination. As these characteristics are available at registration, the proposed association can be evaluated using pre-enrollment trial information. Therefore, we evaluated 17,537 completed interventional clinical trials registered on ClinicalTrials.gov between 2007 and 2024 and sponsored by the 20 largest global pharmaceutical companies. In addition, we examined whether OCI was associated with trial dissemination and if the dissemination varied across levels of operational complexity within sponsor portfolios. Moreover, we evaluated time to dissemination across levels of operational complexity. The potential research investment linked with non-disseminated trials was also evaluated using benchmark costing assumptions. Taken together, this study evaluated if registration-time characteristics are associated with subsequent dissemination and whether they may help identify heterogeneity in dissemination risk within sponsor portfolios.

## Methods

### Study Design and Reporting

We conducted a retrospective observational cohort study to assess whether operational complexity at registration time was linked to the subsequent likelihood and timing of dissemination. This study was reported in accordance with the STROBE guidelines and the RECORD extension for studies using routinely collected health data(12). As the study used de-identified, publicly available registry data, institutional review board approval was not required

### Data Source

All data were obtained from the Aggregate Analysis of ClinicalTrials.gov (AACT) database(13), on 01^st^ February 2026. The AACT is a publicly relational database created by the Clinical Trials Transformation Initiative (CTTI)(14) to provide nightly snapshots of the ClinicalTrials.gov registry. For reproducibility, all analyses were performed through a static AACT snapshot rather than live database. Core trial features, including planned enrollment, number of participating sites, number of countries, clinical phase, therapeutic area, randomization status, masking status, primary completion date, and sponsor identity, were extracted from the AACT studies, sponsors, facilities, countries, and browse_conditions tables using SQL.

### Cohort Selection

The source population consisted of interventional clinical trials registered on ClinicalTrials.gov at or after January 1st, 2007, corresponding to the FDAAA era. To focus on large, resource-rich sponsors with sustained activity across the study period, the cohort was limited to trials sponsored by the 20 largest global pharmaceutical companies according to 2013 global revenue rankings. The year 2013 was chosen as the reference point for this ranking as it falls near the midpoint of the study period and was intended to identify sponsors that were prominent across the FDAAA era.

Due to mergers, acquisitions, and regional subsidiaries, sponsor names were harmonized using explicit string-matching rules. For instance, Janssen and Actelion were mapped to Johnson & Johnson; Allergan was mapped to AbbVie, and Genentech and Chugai were mapped to Roche/Genentech. Additional details of the sponsor harmonization processes were provided in Supplementary Information SI.1.

The primary analytic cohort was restricted to trials with registry status listed as ‘Completed’. Trials with registry statuses of “Terminated”, “Withdrawn” or “Suspended” were excluded as dissemination processes may differ for trials that do not reach completion. A secondary sensitivity cohort required at least 24 months between the primary completion date and the AACT snapshot date. This sensitivity analysis was developed to reduce the misclassification of trials that were still within plausible reporting or publication windows.

### Exposure: Operational Complexity Index (OCI)

We created the Operational Complexity Index (OCI) to quantify trial complexity at registration. The OCI was built on three registration-time trial characteristics available in AACT: (a) planned participant enrollment, (b) number of sites, and (c) number of countries. As these variables were right-skewed, each was transformed using log(x + 1) before analysis. Then, we applied principal component analysis (PCA) to the standardized log-transformed variables to derive a common latent dimension of operational complexity. The first principal component explained 82.4 % of the variance and had positive loadings for each of the aspects for all three variables (log-enrollment: 0.54; log-sites: 0.60; log-countries: 0.59; Cronbach’s alpha = 0.83)(15). Standardized scores from the first principal component were used as the OCI, expressed as standard deviation units. Variance inflation factors were all below 2.5, indicating limited multicollinearity among the three OCI components, as in Table S16(16).

### Outcome Measures

Our primary outcome was trial dissemination status, categorized as disseminated or (non-disseminated) silent. A trial was considered disseminated if results were identified through either of two pathways: (1) posting of summary results to the ClinicalTrials.gov registry results database or (2) a peer-reviewed publication linked to the trial’s NCT identifier via PubMed through the AACT PubMed links table. Trials with no evidence of dissemination through either pathway were classified as non-disseminated/silent. We assessed the reliability of our automated classification pipeline using a blinded manual validation study of 344 randomly selected trials, stratified by algorithmic output and OCI tertile. Blinded reviewers searched PubMed for dissemination evidence using a prespecified hierarchical search protocol. We present full details of our validation procedures and the resulting diagnostic confusion matrix in SI.2.

Our secondary outcome was time to dissemination, which is the number of days from the primary completion date to the earliest dissemination event, either with registry results posting or linked PubMed publications. We censored trials that did not have an identifiable dissemination event by the end of the observation period at the date of the AACT database snapshot.

### Covariates

Multivariable models adjusted for trial-level characteristics associated with dissemination. Covariates included clinical phase (Early Phase 1, Phase 1, Phase 1/2, Phase 2, Phase 2/3, Phase 3, Phase 4, and Unknown), therapeutic area (derived by mapping AACT MeSH browse condition terms to standardized clinical categories (e.g., neoplasms -> Oncology; virus diseases -> Infectious Disease)), randomization status (Randomized vs. Non-randomized), blinding/masking design (Open Label, Single, Double, etc.), and a binary indicator for the FDAAA regulatory era (trial start year < 2008 vs. >= 2008). Sponsor identity was also included as a categorical variable in primary models to account for differences in reporting practices and portfolio composition across sponsors.

### Statistical Analysis

Descriptive statistics were used to summarize cohort characteristics. Continuous variables are reported as median (interquartile range, IQR) and categorical variables are presented as counts/percentages.

Multivariate logistic regression was used to evaluate the association between OCI and dissemination status. Adjusted odds ratios (aOR) and 95% confidence intervals were calculated for OCI and all covariates. To adjust for potential within-sponsor correlation, sponsor-level cluster-robust standard errors were used in logistic regression models. For the time-to-event analysis, Cox proportional hazards(17) models were used to evaluate the association between OCI and timing of dissemination, with hazard ratios (and corresponding 95% CIs) reported for each covariate. Scaled Schoenfeld residual tests(18) were used to assess the proportionality of hazards for each covariate. Because hazard ratios do not directly convey absolute differences in time, the restricted mean survival time (RMST)(19) analysis was also performed. RMST was estimated as the area under the Kaplan-Meier curve up to 1095 days (approximately 3 years) and used to compare dissemination timing between trials in the highest and lowest OCI tertiles.

### Predictive Modeling

Ridge-regularized logistic regression (L2 regularization)(20) was used to evaluate whether non-dissemination could be predicted from registration-time trial characteristics. Categorical variables were one-hot encoded before model fitting. The L2 regularization parameter (λ) was selected through exhaustive grid search. A structural model included registration-time trial features only (OCI, clinical phase, and trial design attributes), and a full model additionally included sponsor identity as categorical indicators. Model discrimination was assessed using the area under the receiver operating characteristic curve (AUC), and calibration was assessed using the Brier score, with both metrics evaluated via 5-fold cross-validation and verified on a hold-out test set.

### Economic Burden Estimation

The economic burden analysis estimated the sunk participant-level research investment associated with non-disseminated trials. For each non-disseminated/silent trial, registered enrollment was multiplied by published phase-specific benchmarks for the per-participant clinical trial costs (SI.5). These estimates should be interpreted as approximate lower-bound benchmarks and do not account for inflation, administrative overhead, or variation in costs across therapeutic areas.

### Sensitivity and Robustness Analyses

Three pre-specified sensitivity analyses were conducted. First, phase-stratified logistic regression models were fitted to assess if the OCI-dissemination association was consistent across clinical development phases (Phase 1, Phase 2, Phase 3). Second, the 24-month sensitivity cohort was analyzed to reduce potential misclassification of trials that remained within plausible reporting windows. Third, a probabilistic bias analysis (PBA)(21) was performed to evaluate the potential effects of false-negative misclassification in the automated dissemination detection pipeline. Misclassification parameters (false negative rate = 10.1%) were used to stimulate corrected OCI-dissemination associations under plausible misclassification scenarios. All analyses were run using Python 3.9 with the pandas, statsmodels, lifelines, and scikit-learn libraries(22, 23). The entire reproducible analysis pipeline (master_pipeline.py) is publicly available in the supplementary repository.

## Results

### Baseline Characteristics and Automated Pipeline Validation

Since 2007, over 212,000 clinical trials have been registered on ClinicalTrials.gov. Of these, 34,741 were interventional studies that were linked to the 20 largest pharmaceutical companies globally, as in Fig 1A. After excluding trials that were not completed, the final analytic cohort comprised 17,537 completed interventional trials sponsored by these 20 companies (Table S1). Only 71.4% of trials (n = 12,518) were classified as being disseminated via at least one pathway, while 28.6% (n = 5,019) were completely silent. Posting registry results was the most common method of dissemination, representing 60.6% of all trials (n = 10,633). Approximately 85.5% of trials (n = 14,995) were initiated during the post-FDAAA era. A large proportion of trials were distributed across the two early phases (Phase 1, 39.4%; n = 6,916; Phase 3, 26.1%; n = 4,584), while a relatively small proportion were distributed across the later phases (Phase 2, 19.6%; n = 3,430; Phase 4, 8.7%; n = 1,529). The planned enrollment was strongly right skewed, with a median of 79 participants (interquartile range [IQR]: 32-274). The standardized OCI across the entire cohort had a mean of 0.00 (standard deviation [SD] = 0.88) and reflected a wide variation in the operational capacity of the trials included in this study (Tables S1 and S11).

**Fig 1.**
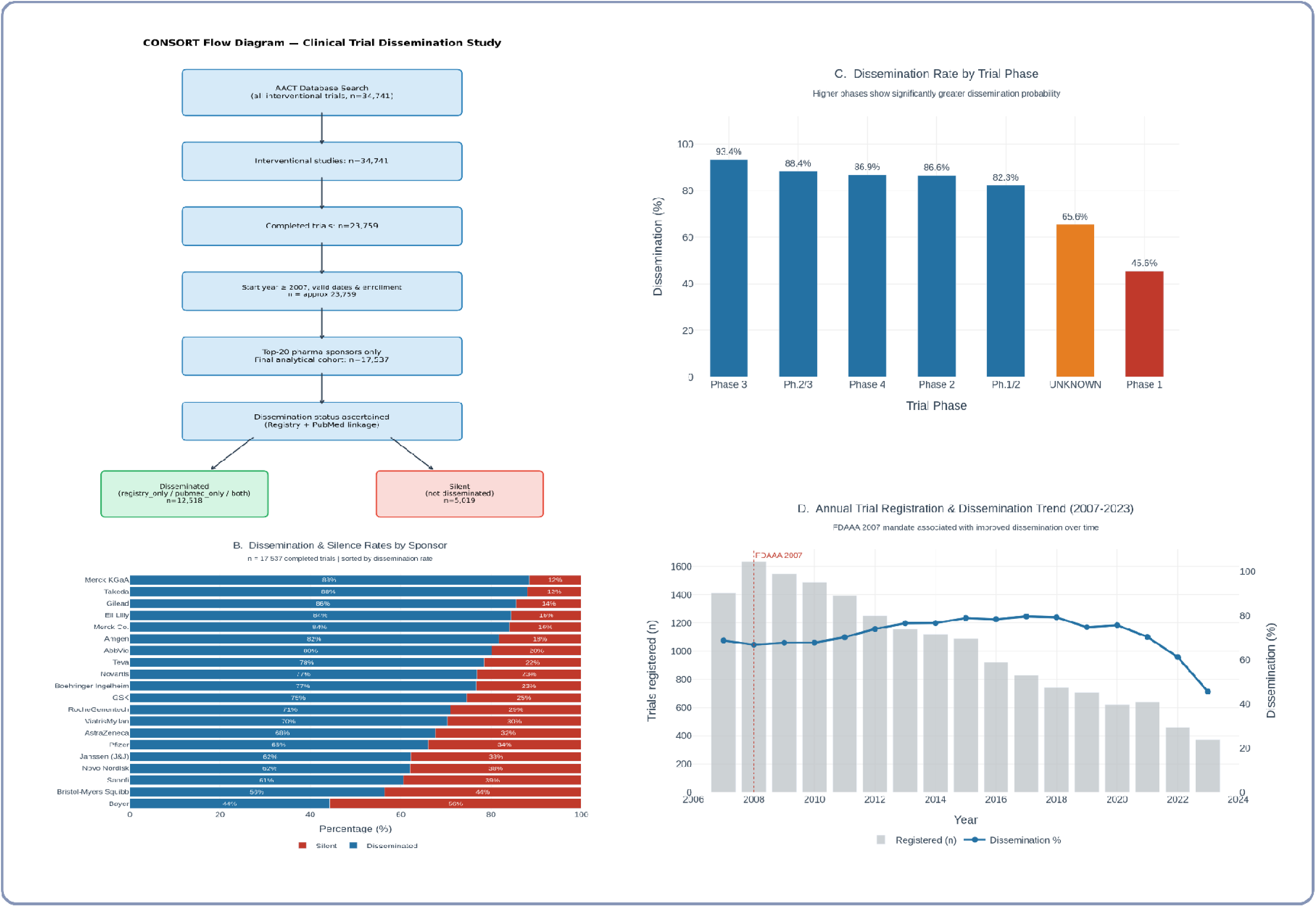
Study cohort and baseline dissemination patterns. (A) CONSORT-style flow diagram showing cohort derivation from 34,741 industry-sponsored interventional trials registered on ClinicalTrials.gov (2007–2024) across the 20 largest pharmaceutical sponsors. After eligibility filtering, 17,537 completed trials formed the final analytic cohort (12,518 disseminated; 5,019 silent). (B) Stacked bar chart showing dissemination (blue) versus silence (red) rates by sponsor among completed trials (n = 17,537). (C) Dissemination rate by trial phase. Dissemination rates varied by development phase, with higher reporting in later-phase trials (Phase 3: 93.4%, Phase 4: 86.9%) compared with early-phase trials (Phase 1: 45.6%). (D) Annual trends in trial registration and dissemination (2007–2023). Bars represent the number of trials registered annually, and the line indicates the dissemination rate. The vertical dashed line marks the FDA Amendments Act (2007). Dissemination rates generally increased after the mandate but declined in recent years.

The automated dissemination classification pipeline performed well in a blinded manual validation of a stratified random sample of 344 trials (Table S12). Overall accuracy of the pipeline was 92.1%, with a sensitivity of 89.8%, specificity of 96.1%, and Cohen’s Kappa (κ) of 0.84. There was no statistically significant difference in the agreement of the pipeline and the gold standard across the three OCI tertiles (low: 91.3%, κ = 0.82; medium: 92.2%, κ = 0.84; high: 92.9%, κ = 0.86). The false negative rate of 10.1% was primarily due to events of dissemination that occurred through abstracts at conferences or proprietary sponsor repositories (SI.2).

### Sponsor-Level Dissemination Patterns

Dissemination rates varied across the 20 largest pharmaceutical sponsors, ranging from 44.3% (Bayer) to 88.4% (Merck KGaA) (Fig 1B, Table S11). Among high-volume sponsors, GSK had a dissemination rate of 74.6% and contributed 515 silent trials, the largest absolute count in the cohort. Janssen (J&J) and Pfizer contributed 521 and 572 silent trials, respectively, despite aggregate dissemination rates of 62.2% and 66.0%. Among high-volume sponsors, Takeda (88.0%), Eli Lilly (84.4%), and Merck Co. (84.1%) had the highest aggregate dissemination rates (Table 1, Table S11). Mean sponsor-level OCI values ranged from −0.31 (Merck Co.) to +0.41 (Teva), indicating heterogeneity in the operational scale of sponsor portfolios (Table S11). Annual trial initiation data showed that all 20 sponsors registered trials throughout the 2007-2024 study period (Table S2).

**Table 1.**
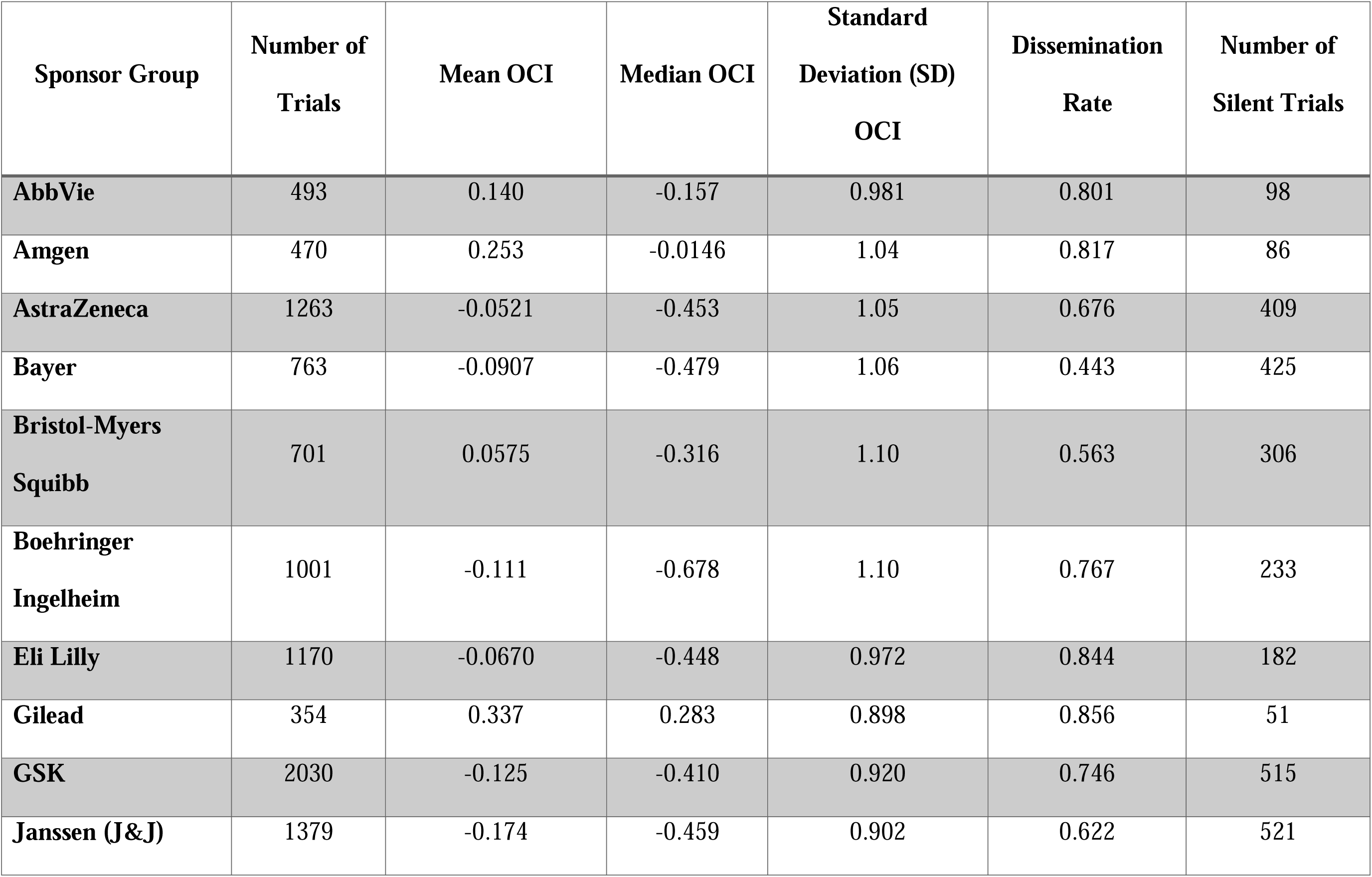
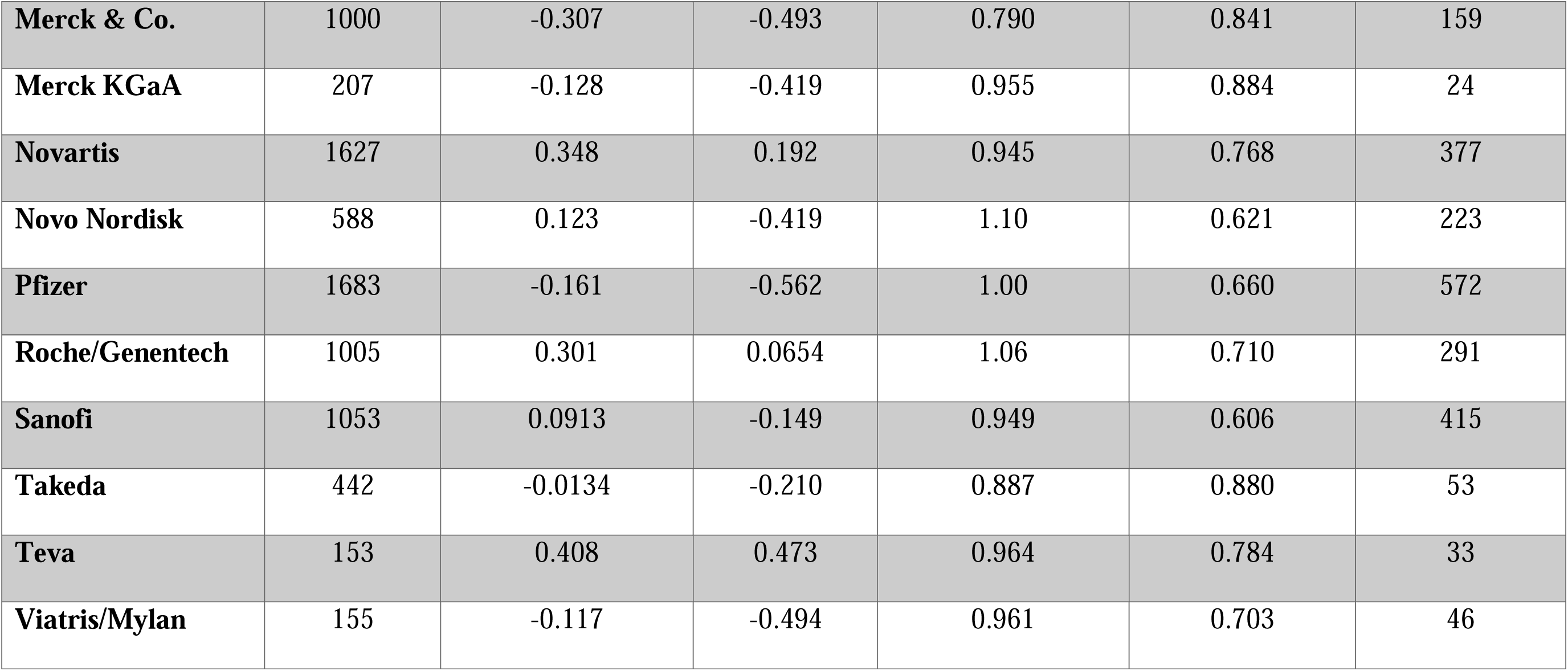
Characteristics of 17,537 completed clinical trials by pharmaceutical sponsor.

| **Sponsor Group** | **Number of Trials** | **Mean OCI** | **Median OCI** | **Standard Deviation (SD) OCI** | **Dissemination Rate** | **Number of Silent Trials** |
| --- | --- | --- | --- | --- | --- | --- |
| **AbbVie** | 493 | 0.140 | -0.157 | 0.981 | 0.801 | 98 |
| **Amgen** | 470 | 0.253 | -0.0146 | 1.04 | 0.817 | 86 |
| **AstraZeneca** | 1263 | -0.0521 | -0.453 | 1.05 | 0.676 | 409 |
| **Bayer** | 763 | -0.0907 | -0.479 | 1.06 | 0.443 | 425 |
| **Bristol-Myers Squibb** | 701 | 0.0575 | -0.316 | 1.10 | 0.563 | 306 |
| **Boehringer Ingelheim** | 1001 | -0.111 | -0.678 | 1.10 | 0.767 | 233 |
| **Eli Lilly** | 1170 | -0.0670 | -0.448 | 0.972 | 0.844 | 182 |
| **Gilead** | 354 | 0.337 | 0.283 | 0.898 | 0.856 | 51 |
| **GSK** | 2030 | -0.125 | -0.410 | 0.920 | 0.746 | 515 |
| **Janssen (J&J)** | 1379 | -0.174 | -0.459 | 0.902 | 0.622 | 521 |
| **Merck & Co.** | 1000 | -0.307 | -0.493 | 0.790 | 0.841 | 159 |
| **Merck KGaA** | 207 | -0.128 | -0.419 | 0.955 | 0.884 | 24 |
| **Novartis** | 1627 | 0.348 | 0.192 | 0.945 | 0.768 | 377 |
| **Novo Nordisk** | 588 | 0.123 | -0.419 | 1.10 | 0.621 | 223 |
| **Pfizer** | 1683 | -0.161 | -0.562 | 1.00 | 0.660 | 572 |
| **Roche/Genentech** | 1005 | 0.301 | 0.0654 | 1.06 | 0.710 | 291 |
| **Sanofi** | 1053 | 0.0913 | -0.149 | 0.949 | 0.606 | 415 |
| **Takeda** | 442 | -0.0134 | -0.210 | 0.887 | 0.880 | 53 |
| **Teva** | 153 | 0.408 | 0.473 | 0.964 | 0.784 | 33 |
| **Viatris/Mylan** | 155 | -0.117 | -0.494 | 0.961 | 0.703 | 46 |

### Operational Complexity and Dissemination Probability

Higher OCI was associated with greater odds of trial dissemination. Specifically, each 1-SD increase in OCI was associated with higher odds of dissemination (adjusted OR of 2.40, 95% CI: 2.23–2.60; p < .001; Table 2, Table S4). Dissemination rate increased across higher levels of OCI within sponsor portfolios (Fig 2A, Fig S5). For instance, dissemination ranged from 47 % among trials in the lowest OCI decile to 95 % among those in the highest OCI decile (Fig 2A, Fig S5). Coming to the sponsor level, mean OCI was positively correlated with overall dissemination rate (Pearson Correlation Coefficient r = 0.75, p < 0.001; Fig 2B). Among sponsors, the mean difference in dissemination between low- and high-complexity trials was 40 % points (range: 4–70 % points; p < 0.001 for heterogeneity among the 20 sponsors). All 20 sponsors showed a positive OCI-dissemination gradient, and none showed a statistically significant negative association. Overall, these findings suggest that the OCI-dissemination association was consistent across sponsors rather than being driven by a single company.

**Figure 2.**
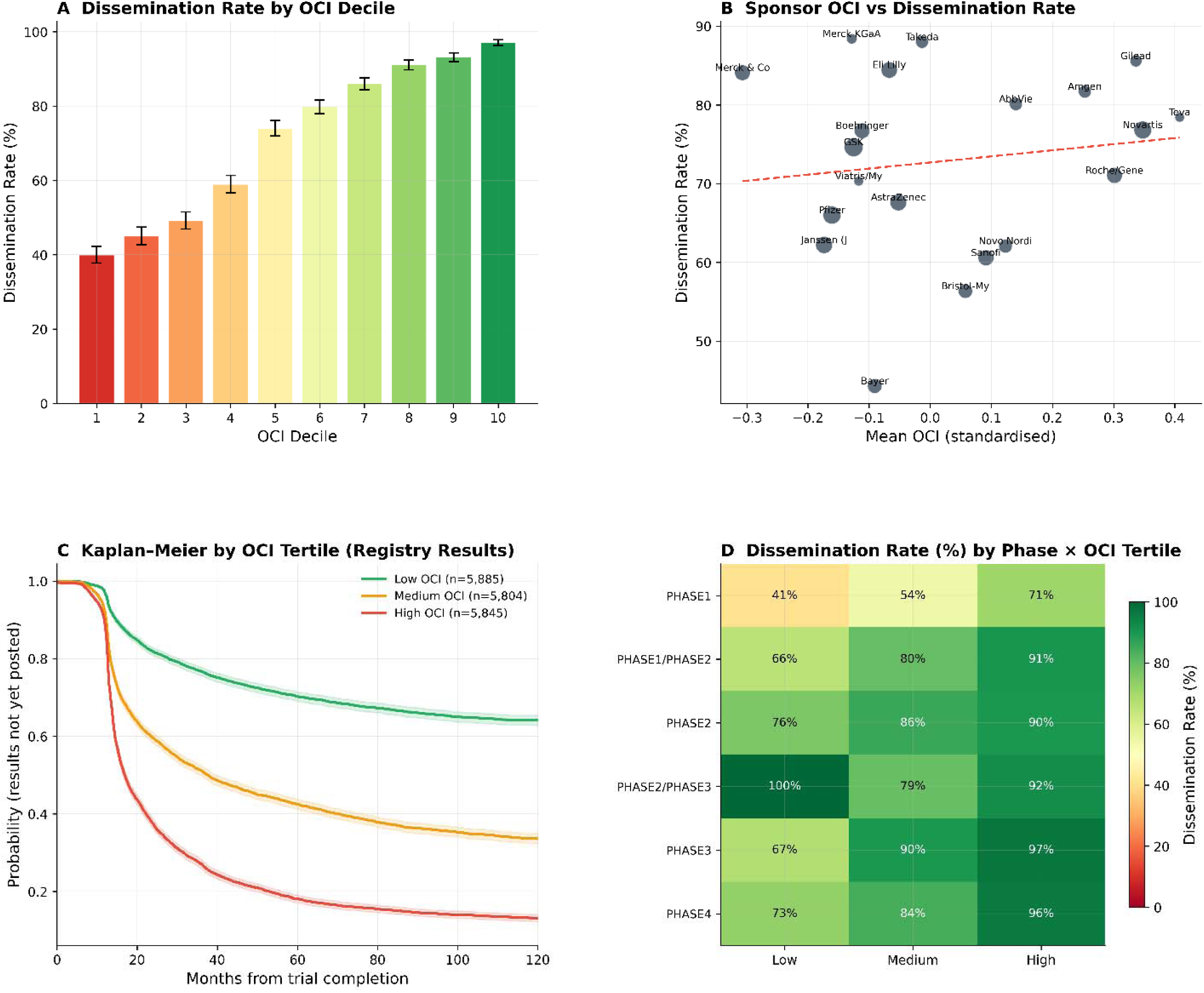
Operational complexity dose-response relationship with clinical trial dissemination. **(A)** Dissemination rates increase monotonically cross Operational Complexity Index (O I) deciles, from ≈47% (lowest decile) to ≈95% (highest decile; n=17,537 completed trials). **(B)** Dissemination rate regresses linearly against mean standardized OCI per sponsor (R²≈0.75; 20 sponsors plus “Other”), confirming sponsor-level gradient. **(C)** Kaplan–Meier curves stratified by OCI tertilesm (low/medium/high), showing high-OCI trials disseminate si nificantly faster (restricted mean surviv l time difference ≈310.88 days over 3 y ars). **(D)** Heatmap of dissemination rates by clinical trial phase and OCI tertile, demonstrating persistence across all ph ses (Phase 1: 47–53%; Phase 4: 71–94%). Data derived from TableS4_Sponsor_OCI_ tats.csv and primary survival analyse .

**Table 2.**
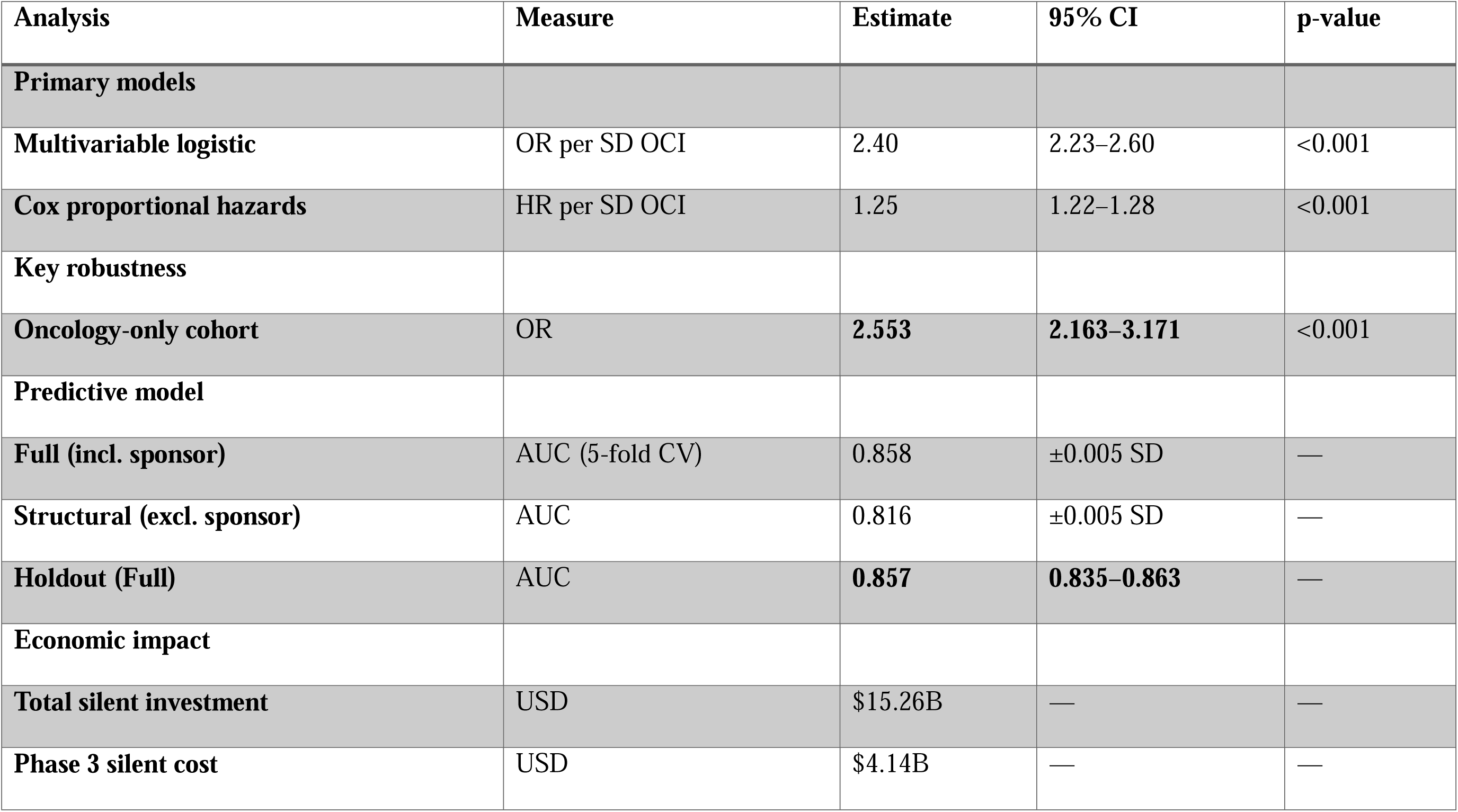
Core associations between operational complexity and dissemination.

Clinical trial phase was also associated with dissemination after adjustment for OCI and other covariates (Table S4). Compared to Phase 1 trials, Phase 3 and Phase 4 trials had significantly higher odds of being disseminated (Phase 3: aOR = 13.51, 95% CI: 5.81–31.43, p < 0.001; Phase 4: aOR = 12.16, 95% CI: 5.21–28.40, p < 0.001). Trials initiated in the post-FDAAA era also had higher odds of dissemination (aOR = 2.00, 95% CI: 1.78–2.25, p < 0.001). Through therapeutic area, oncology and infectious disease trials had higher odds of dissemination (Oncology: aOR = 1.57, 95% CI: 1.25–1.97, p < 0.001; Infectious Disease: aOR = 1.45, 95% CI: 1.14–1.84, p = 0.002) (Table S4). The phase-by-OCI tertile heatmap showed that the OCI gradient in dissemination was present across clinical phases (Fig 2D). For example, within the Phase 4 trials, the dissemination rate varied from 71% within the low-OCI tertile to 94% within the high-OCI tertile.

### Temporal Dynamics and Time-to-Dissemination

High OCI was associated with earlier dissemination in Cox proportional hazards models. Each 1 SD increase in OCI was associated with a higher hazard of dissemination (HR 1.25, 95 % CI 1.22 – 1.28; p < 0.001; Table 2). Kaplan-Meier plots stratified by OCI tertile showed clear separation in dissemination trajectories across follow-up (Fig 2C; Figs S4 and S6) (24). Using RMST over 1.095 days (3 years), high-OCI trials were disseminated a mean of 310.88 days earlier than low-OCI trials (RMST 659.10 days vs 969.98 days; difference −310.88 days, 95% CI −320.96 to −300.59; p<0.001; Table S6). Trials initiated in the post-FDAAA era also had a higher hazard of dissemination than trials initiated before 2008 (HR 1.83; 95% CI 1.73 – 1.94; p < 0.001), indicating earlier dissemination in adjusted analysis. At the Sponsor level, Merck Co., (HR 1.97), Merck KGaA (HR = 1.88), Boehringer Ingelheim (HR 1.77) and Eli Lilly (HR 1.75) were tethered to earlier dissemination, whereas Bayer (HR = 0.28), Sanofi (HR = 0.37), and Roche/Genentech (HR = 0.63) were associated with later dissemination (Table S5).

### Predictive Modeling

The structural ridge-regularized logistic regression model trained using registration-time variables only (clinical phase, trial design, and OCI), achieved a mean cross-validated AUC of 0.816 (SD 0.005), with holdout AUC of 0.814 and Brier score of 0.152 (Tables S7; Figs S1 and S2). (Tables S7; Figs S1 & S2). Adding sponsor identity as categorical indicators increased model discrimination, yielding a mean cross-validated AUC of 0.858 (SD 0.005), holdout AUC of 0.857, and Brier score of 0.134 (Table S7). For comparison, a phase-only model achieved an AUC of 0.776 was obtained with a Phase-only model and 0.757 was obtained with an Enrollment-only model. This confirms that the composite OCI is providing substantial predictive value beyond either of these individual features, i.e., increasing AUC by 0.038 above Phase-only and 0.057 above Enrollment-only (Table S7).

Models of each component (enrollment, site or country) used individually resulted in equivalent AUC values (hold-out AUC = 0.796; 95% CI:0.782-0.809; Table S17) which supports the use of the three variables as surrogate measures of a single structural dimension and thus supports the use of a composite measure derived through PCA rather than independent predictors.

Using a decision threshold of 0.30, the full model had a sensitivity of 0.798 and specificity of 0.748 (PPV = 0.559; NPV = 0.902) and correctly identifies about 80% of the trials that will be unreported with an acceptable false-positive rate. Using decision curve analysis, it is clear that the full model has positive net benefit throughout the clinically relevant range of thresholds (0.25 – 0.35) and clearly outperforms the ’flag all trials’ and simpler model specification approaches (Fig S12).

Additionally, the Brier score of the full model (0.134) is 34% better than the null model Brier score (0.204) and indicates good probabilistic calibration at all levels of risk. Inspection of the standardized coefficients from the model indicate that the OCI is the largest predictor of the probability of a trial being disseminated, followed by clinical phase and specific trial design attributes. Sponsorship is the next largest contributor of predictive information after the OCI (Table S4).

### Economic Burden of Non-Dissemination

Using phase-specific per-participant cost benchmarks (SI.5), the 5,019 non-disseminated trials corresponded to a primary estimated US$15.26 billion in sunk research investment (Tables S8, Fig S3). Estimated costs varied across development phases. The 302 silent Phase 3 trials accounted for an estimated US $4.14 billion, whereas the 3,762 silent phase 1 trials accounted for an estimated US$3.26 billion. The 459 silent Phase 2 trials contributed an estimated $2.51 billion. An additional 200 silent trials with uncertain phase classification were assigned an estimated S$4.32 billion under the primary assumption that Phase 1 benchmark costs (US$20,000 per participant) applied (SI.5). Sensitivity analyses for these 200 trials evaluated alternative enrollment assumptions, including capping enrollment at the 99th cohort percentile, median imputation, and exclusion of enrollment values. Under these alternative assumptions, the estimated total ranged from US$10.94 billion to US$11.41 billion, indicating substantial sensitivity of the cost estimate to phase-classification and enrollment assumptions (Table S24). In absolute terms, estimated sponsor-level burden was highest for Roche/Genentech (US$3.52 billion), Sanofi (US$2.08 billion), and Novartis (US$1.78 billion), and lowest for Merck KGaA (US$0.02 billion), Viatris/Mylan (US$0.05 billion), and Gilead (US$0.12 billion) (Fig 3B; Table S9).

**Fig 3.**
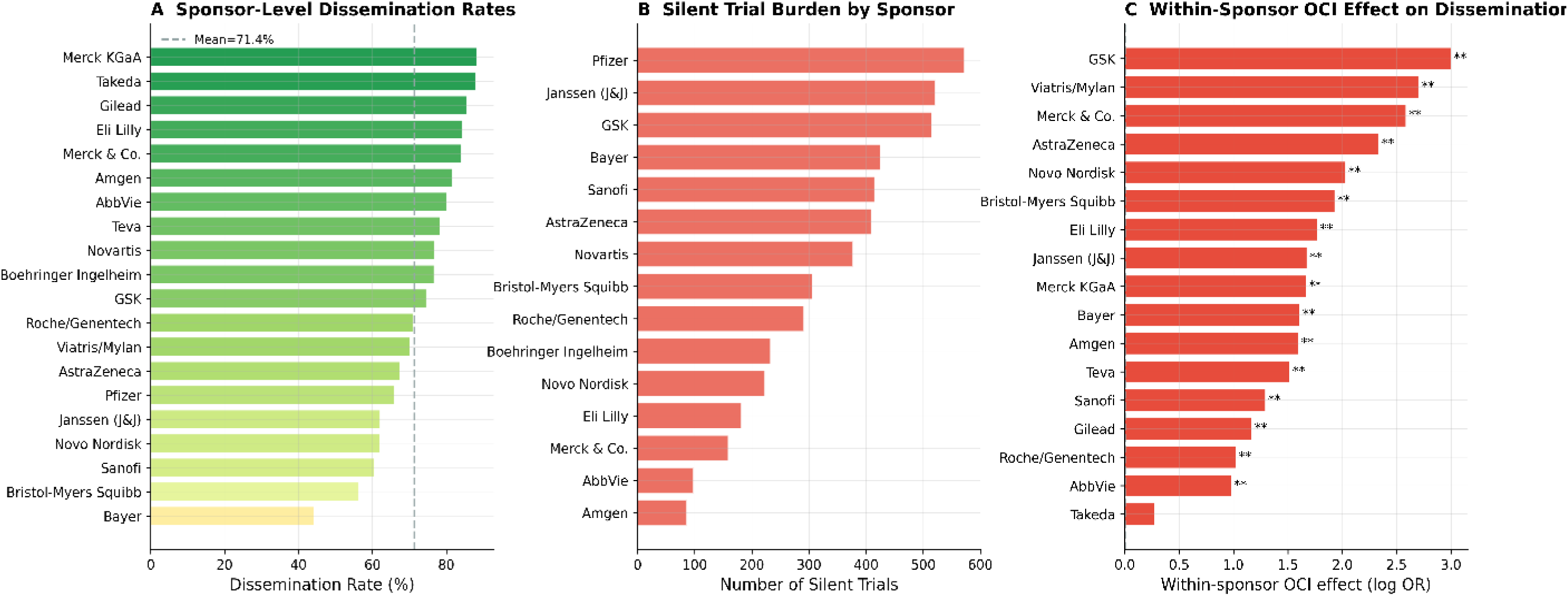
Within-sponsor accountability gaps and economic burden of selective trial transparency. **(A)** Dissemination rates by pharmaceutical sponsor (range 44–88%; n=17,537 completed trials; data from Table S4). **(B)** Economic burden measured as number of silent trials by sponsor (5,019 total; Roche, Sanofi, Novartis contribute largest absolute burden). **(C)** Accountability gaps defined as the percentage point difference in dissemination rates between high- versus low-OCI tertile trials *within each individual sponsor’s portfolio* (mean gap = 40 percentage points; range 4–70 pp; all sponsors exhibit positive OCI gradients, p<0.001 for heterogeneity). The 5,019 silent trials across all sponsors represent an estimated **$15.26B** in undisseminated participant-borne research investment.

### Robustness and Sensitivity Analyses

The positive OCI-dissemination association was consistent across sensitivity analyses. In phase-specific logistic regression models, OCI remained positively associated with dissemination in Phase 1 (aOR = 2.40, p < 0.001); Phase 2 (aOR = 1.57, p < 0.001); and Phase 3 (aOR = 2.94, p < 0.001) (Table S13). These findings suggest that the OCI-dissemination association was not explained solely by the clinical phase. In the 24-month follow-up sensitivity cohort, which restricted analyses to trials with longer elapsed time since primary completion, the association was stronger than in the primary cohort (aOR 4.56 vs 2.40; p<0.001; Table S13).

When analyses were restricted to oncology trials, OCI remained positively associated with dissemination (OR = 1.59, p < 1 × 10^−58^; Table 2). This finding reduces concern that the primary association was driven solely by differences across therapeutic areas. Probabilistic bias analysis using the 10.1% false-negative misclassification rate from manual validation yielded a corrected OCI–dissemination association that remained positive (simulated aOR 4.19, 95% CI 4.07–4.30) under plausible misclassification scenarios. The larger corrected estimate relative to the primary model (aOR 4.19 vs 2.40) suggests that false-negative outcome misclassification may have attenuated the primary association (Table S14). Multicollinearity among OCI components was low, with all variance inflation factors below 2.5 (log enrollment 1.88, log sites 2.31, log countries 2.24; Table S16).

Individual component models showed similar predictive performance (AUC 0.796 for each), consistent with these variables capturing a shared structural dimension. Principal Components Analysis also supported the use of OCI as a summary measure of operational scale (Table S15). (25) The first principal component explained 82.4% of the variance among the three OCI components, with positive loadings for all (0.54 – 0.60; Cronbach’s alpha = 0.83).

To examine whether the OCI association persisted for a dissemination pathway less dependent on journal publication, we evaluated registry results posting separately. The OCI gradient persisted for registry results posting (aOR 3.46, 95% CI 3.30–3.63; p<0.001). Posting rates increased from 32.6% (1,909/5,848) in the lowest OCI tertile to 85.6% (5,003/5,846) in the highest OCI tertile (Table S24; Fig S11), suggesting that the primary association was not limited to publication-linked dissemination pathways.

## Discussion

### Summary of findings

In this retrospective cohort of 17,537 completed interventional clinical trials sponsored by the 20 largest global pharmaceutical companies, operational scale at registration, quantified using OCI, was consistently associated with subsequent dissemination. Each 1-SD increase in OCI was associated with a higher odd of dissemination (aOR = 2.40; 95% CI: 2.23–2.60; p < 0.001), with dissemination rates increasing from 47% in the lowest OCI decile to 95% in the highest. This gradient was observed across sponsors, clinical phases, and therapeutic areas. Higher-complexity trials were also disseminated earlier than lower-complexity trials, with an RMST difference of 310.88 days (95% CI: 300.59–320.96; p < 0.001). Registration-time trial characteristics also showed predictive signal for non-dissemination, with a ridge-regularized model achieving an AUC of 0.816 using structural features alone and 0.858 when sponsor identity was included. Using benchmark phase-specific costing assumptions, the 5,019 non-disseminated trials corresponded to an estimated US$10.9–15.3 billion in sunk research investment, although this estimate was sensitive to assumptions regarding phase classification and enrollment (Table S24). Taken together, these findings suggest that non-dissemination is unevenly distributed across trial types and is associated with registration-time operational characteristics.

### The Structural Shadow: A New Framework for Understanding Selective Transparency

These findings suggest that smaller trials with lower operational visibility were less likely to be disseminated. Prior research has often reported aggregate rates of non-reporting across sponsors and phases, implicitly treating non-dissemination as relatively uniform within sponsor portfolios(8). Our findings suggest that this characterization may overlook importance within-sponsor heterogeneity. Notably, even among sponsors with high aggregate dissemination rates, such as Takeda (88.0%) and Eli Lilly (84.4%), lower-OCI trials were less likely to be disseminated than higher-OCI trials. These findings suggest that aggregate compliance metrics may be insufficient to detect within-sponsor heterogeneity in dissemination patterns(26). In other words, a sponsor may perform well on aggregate reporting measures while still showing lower dissemination among smaller trials with lower operational visibility.

OCI captures three related dimensions of operational scale: planned enrollment, number of facilities, and number of countries. Principal component analysis supported combining these variables into a single summary measure (loadings 0.54–0.60; variance explained 82.4%; Cronbach _α_=0.83). Model comparisons also suggested that OCI provided better discrimination than enrollment alone (AUC 0.814 versus 0.757), supporting the added value of a multidimensional measure of operational scale. The somewhat higher loadings for facility and country count may indicate that geographic dispersion contributes additional information beyond enrollment size alone.

There are several plausible explanations for the gradient observed in the data. For example, large multicenter clinical trials attract multiple institutional actors including, academic investigators, patient advocacy groups, government regulators, etc., who each provide independent institutional pressures to report results(10). Conversely, smaller Phase I pharmacokinetic or safety studies often result in results being reported only internally to the regulatory agencies and sponsors(27). Such trials may also be more likely to yield null or inconclusive findings, which could reduce publication likelihood, although these mechanisms were not directly examined in this study(28). Publication bias favoring larger studies or positive findings could also contribute to the observed pattern. The probabilistic bias analysis further suggested that false-negative outcome misclassification may have attenuated the primary association, with a corrected adjusted odds ratio of 4.19 (95% CI 4.07–4.30) compared with 2.40 in the primary model.

### Participant Contributions and the Ethics of Sunk Investment

These findings also have ethical and public health implications that warrant careful consideration. Using benchmark participant-level trial cost assumptions, we estimated that 5,019 non-disseminated trials in this cohort were associated with substantial sunk research investment. The estimated burden ranged from US$10.9 billion to US$15.3 billion, with the primary estimate of US$15.26 billion being sensitive to assumptions regarding phase classification and enrollment (Table S24). By phase, the estimated burden included US$4.14 billion from 302 non-disseminated Phase 3 trials, US$3.26 billion from 3,762 non-disseminated Phase 1 trials, and US$2.51 billion from 459 non-disseminated Phase 2 trials. These estimates reflect research investment associated with findings that were not captured through prespecified public dissemination pathways; they do not imply that such trials lacked internal scientific or regulatory value. However, when findings are not publicly disseminated, their contribution to the broader scientific literature and evidence base is reduced. International ethics guidance emphasizes public dissemination of research results, including the principles articulated in the Declaration of Helsinki. The concentration of non-dissemination among lower-complexity trials suggests that current transparency frameworks may not capture dissemination risk uniformly across trial portfolios.

### Policy Implications: From Reactive Monitoring to Proactive Surveillance

The predictive modeling results suggest that non-dissemination risk may be partially identifiable at registration. A model based on registration-time features alone, including OCI, clinical phase, and trial design characteristics, achieved an AUC of 0.816. This level of discrimination suggests that registration-time risk stratification may warrant further evaluation in future transparency-monitoring research. If prospectively validated, such approaches might support targeted monitoring, reminders, or additional follow-up for trials at higher predicted risk of non-dissemination. More broadly, these findings raise the possibility that transparency oversight could eventually incorporate prospective risk assessment in addition to retrospective compliance monitoring. We also found that the post-FDAAA era was associated with earlier dissemination (HR 1.83, 95% CI 1.73–1.94; p<0.001), consistent with an association between regulatory context and dissemination timing. However, this observational analysis does not establish the effect of any specific policy mechanism.

### Limitations

This study has several limitations. First, the automated dissemination-classification pipeline had a false-negative rate of 10.1%, indicating that some trials classified as non-disseminated may have been disseminated through channels not captured by the prespecified ascertainment strategy, such as conference abstracts, sponsor portals, or grey literature. Conference abstracts do not necessarily meet full reporting standards, and the implications of this definitional choice for validation performance are described in SI.2. In probabilistic bias analysis, incorporating this false-negative rate yielded a corrected association estimate of aOR 4.19 (95% CI 4.07–4.30), compared with 2.40 in the primary model. This suggests that false-negative outcome misclassification may have attenuated the primary association.

Second, the economic estimates represent approximate benchmark-based participant-level sunk investment costs derived from registered enrollment and phase-specific per-participant cost assumptions. These estimates do not include trial-specific administrative costs, inflation adjustments, or variation in cost structures across therapeutic areas, and should therefore be interpreted as approximate rather than exact financial evaluations. Sensitivity analyses showed that these estimates varied materially according to assumptions regarding uncertain phase classification (Table S24).

Third, the observational design precludes causal inference. Although the OCI–dissemination association was robust across multiple analyses, it should not be interpreted as evidence that operational complexity itself causes dissemination. Unmeasured factors, including trial results, internal sponsor policies, therapeutic-area-specific requirements, and contractual obligations with collaborators, may influence both operational complexity and dissemination. A particular limitation is the lack of trial outcome data. However, the persistence of the OCI gradient for registry results posting (aOR 3.46, 95% CI 3.30–3.63; Table S24; Fig S11) suggests that the observed association was not limited to publication-linked dissemination pathways.

Finally, the cohort was limited to the 20 largest global pharmaceutical companies; generalizability to smaller biotechnology firms, academic medical centers, or government-sponsored trials remains uncertain.

### Conclusion

In this cohort, non-dissemination was unevenly distributed across trial types and was strongly associated with registration-time operational characteristics. Across 17 years of trials sponsored by the 20 largest pharmaceutical companies, greater operational complexity at registration was consistently associated with a higher likelihood of dissemination and earlier dissemination. The within-sponsor gradient, averaging 40 percentage points between the least and most complex trials, suggests that aggregate transparency metrics may obscure meaningful heterogeneity within sponsor portfolios. Because registration-time characteristics showed predictive signal for non-dissemination, future work could evaluate whether prospectively validated monitoring approaches may complement retrospective compliance auditing. The potential scale of non-dissemination identified in this cohort underscores the importance of more complete and timely dissemination of trial results.

## Data Availability

All data produced in the present study are available upon reasonable request to the authors

## Acknowledgement

During the preparation of this work, Grammarly was used to assist with language editing and refinement of the manuscript. After using this tool, the authors reviewed and edited the manuscript as needed and take full responsibility for the content of the **publication.**

## Data Availability

All relevant data underlying the findings of this study are within the manuscript and its Supporting Information files.

