## Supplementary for "Operational complexity predicts selective non-dissemination within pharmaceutical sponsor portfolios: a retrospective cohort study"

**Short title:** *Operational complexity and clinical trial dissemination*

Ahmed M. Sayed[1,2]*, Phan Thieu Huan[2,3], Thuan K. Nguyen [2,4], Eman Fathy [2,5,6], Toka Aziz[2,7], Duong Van Tho[2, 8], Nguyen Tien Huy[9,10,11,]*

**Affiliations**

[1] Department of Chemistry, College of Science, Purdue University, West Lafayette, IN 47907, USA
[2] Online Research Club (www.onlineresearchclub.org), Nagasaki, Japan
[3] University of Medicine and Pharmacy, Hue University, Vietnam
[4] University of Medicine and Pharmacy at Ho Chi Minh City, Ho Chi Minh City 70000, Vietnam
[5] Infection Control and Public Health Administration, Egypt Healthcare Authority (EHA), Egypt.
[6] Faculty of Pharmacy, Beni-Suef University, Egypt.
[7] Faculty of Medicine, Alexandria University, Alexandria, Egypt
[8] Faculty of Medicine and Pharmacy, Binh Duong Economics and Technology University, Vietnam
[9] Institute of Research and Development, Duy Tan University, Da Nang, Vietnam
[10] School of Medicine and Pharmacy, Duy Tan University, Da Nang, Vietnam
[11]School of Tropical Medicine and Global Health (TMGH), Nagasaki University, 1-12-4 Sakamoto, Nagasaki, 852-8523, Japan

Authors' email and ORCID:

| Section | Title | Purpose / Content |
|---|---|---|
| **SI.1** | **Extended Data Source and Sponsor Harmonization Protocols** | **Data source, sponsor harmonization, and cohort assembly framework.** |
| **SI.2** | **Extended Manual Validation Protocol and Diagnostic Confusion Matrix** | **Manual validation design, search protocol, and diagnostic performance of dissemination classification.** |
| **SI.3** | **OCI Component Analysis and Multicollinearity Checks** | **OCI construction, correlations, PCA rationale, and multicollinearity diagnostics.** |
| **SI.4** | **Predictive Machine Learning Pipeline and Hyperparameter Tuning** | **Predictive modeling workflow, feature encoding, tuning, and model evaluation.** |
| **SI.5** | **Economic Burden Cost Inputs and Assumptions** | **Cost assumptions and economic burden estimation framework for silent trials.** |
| **SI.6** | **Sensitivity and Robustness Analyses** | **Sensitivity, robustness, and bias analyses supporting the main findings.** |
| **SI.7** | **Proportional Hazards Diagnostics** | **Schoenfeld diagnostics and rationale for RMST-based interpretation.** |
| **SI.8** | **Analytic Cohort Derivation and Missing Data Handling** | **Narrative cohort derivation, analytic restrictions, and missing-data handling.** |
| **SI.9** | **RECORD / STROBE Checklist Aligned to Manuscript** | **Mapping of reporting checklist items to locations in the manuscript and supplement.** |
| **Supplementary Figs** | **Figs S1–S13** | **Extended visualizations and diagnostic displays supporting the main results.** |

| | | |
|---|---|---|
| **Supplementary Tables** | **Tables S1–S27** | **Extended numerical results, validation outputs, and sensitivity analyses.** |
| **Supplementary References** | **References cited only in the SI** | **References cited exclusively in the Supplementary Information.** |

### SI.1. Extended Data Source and Sponsor Harmonization Protocols

To provide a comprehensive analysis of the large pharmaceutical companies (and thereby assess how these well-established and well-funded firms operated during the full period of FDAAA), the cohort was limited to only the 20 largest pharmaceutical companies in 2013 based on total worldwide revenues. In addition to creating a single entity name to use when analyzing the data from each company, there were also corporate transactions (e.g., mergers/acquisitions) that changed the reporting names of the companies or had regional subsidiary sites report under different names (e.g., Actelion/Janssen - both would be assigned to Johnson & Johnson). Therefore, using an array of explicit string matches, the researchers performed sponsor harmonization to create a uniform naming convention for all study sponsors. For instance, "Allergen" was renamed to "AbbVie", "Actelion" and "Janssen" were renamed to "Johnson & Johnson", and "Genentech" was renamed to "Roche/Genentech". In order to assemble the primary analytical cohort, the researchers used relational SQL queries to extract the foundational data from the AACT database and used the studies, sponsors, facilities, countries, and browse_conditions tables to build the cohort.

### SI.2. Extended Manual Validation Protocol and Diagnostic Confusion Matrix

To prevent a systematic bias in the automatic classification of trial status while linking postings on ClinicalTrials.gov to their corresponding publications indexed in PubMed to their NCT identifiers, a thorough manual validation study was conducted. The study included a stratified random sample of 344 completed clinical trials from the total analytical cohort that was stratified in proportion to the two variables used in the strata: the algorithmic outputs ("Both", "Registry_only", "PubMed_only", "Silent") and the Operational Complexity Index (OCI) into three equal groups (Low, Medium, High). Blind reviewers performed an exhaustive hierarchical search process. First, they manually examined the ClinicalTrials.gov posting to obtain the tabulated results. Second, they utilized standardized Boolean queries in PubMed/MEDLINE and Google Scholar, using the NCT identifier, principal investigator name, and terms related to the intervention and condition to find relevant articles. Third, they searched proprietary corporate transparency databases, such as the GSK Study Register and the Novartis Clinical Trial Results database. Finally, the blind reviewers queried the grey literature of trials classified as "Silent" by the algorithm by searching for conference abstracts and regulatory assessment reports such as the FDA drug approval package. The manual review identified 195 True Positives and 122 True Negatives. There were five false positives, i.e., broken links to unrelated articles, and twenty-two false negatives, all due to the fact that these articles were published solely in non-indexed conference abstracts or proprietary grey literature. In summary, the automated pipeline had high precision with an accuracy of 92.1%, a Cohen's of 0.84, a False Positive Rate of 3.94% and a Positive Predictive Value of 97.5%. The sample size (n = 344) of the validation set was determined to yield a precision of ± 0.05 around the expected Cohen's κ of ~ 0.84, similar to sample size guidelines recommended in agreement studies[1]. These guidelines indicate that 300-400 observations will be sufficient to achieve this level of precision.

#### SI.3. OCI Component Analysis and Multicollinearity Checks

The three key base metrics for the Operational Complexity Index are the planned enrollment of participants for the study, the number of facilities that will be involved in the study, and the number of countries where the study will take place. Due to extreme right skewing of clinical trial structural data, each of these variables had a natural logarithm transformation applied to them before they could be z-score normalized to achieve normal distributions for comparison. Pearson correlation coefficients were used to confirm the structural integrity of the index by assessing the internal relationships between the log transformed components of the OCI and the z-scores for the OCI (Table S3). Variance inflation factors (VIF) were also calculated for all continuous and categorical predictor variables included in the primary multivariable regression model(s) to check for the presence of multicollinearity among the structural predictors. All of the component VIFs were safely less than the accepted threshold of 4, thus confirming there was no evidence of multicollinearity among the structural predictors.

#### SI.4. Predictive Machine Learning Pipeline and Hyperparameter Tuning

The supervised machine learning pipeline with Ridge logistic regression used to predict the likelihood of a trial being silent in terms of outcomes based on baseline trial characteristics was trained using a 5-fold cross validation approach. One hot encoding was utilized to process all categorical features, including Clinical Phase, Sponsor Identity, and Therapeutic Area. To limit overfitting from the very high dimensional space due to the structural features, the L2 regularizer() was tuned by performing a full grid search. Both the "Structural Model", where the model was blinded to Sponsor Identity, and the "Full Model", where the model knew the Sponsor Identity and included the structural features, were evaluated and compared. The "Full Model" had an Area Under the Curve (AUC) of 0.858 while the "Structural Model" had an AUC of 0.816. Additional comparison metrics for each model, including Brier Scores and Hold Out AUCs, can be found in Table S7.

In order to alleviate the reviewer's concern that OCI simply represents trial size, we examined the performance of four different models: the Enrollment Only Model, Sites Only Model, Countries Only Model and the Composite OCI Model (Table S17). Each of the component models produced nearly identical AUC values (0.796) indicating that the three structural features have captured substantially similar predictive information. The rationale for reducing the number of dimensions of these features into a single index through PCA is to retain the ability to discriminate the operational scale of the trials while producing an interpretative and theoretically-based composite measure (PC1=82.4% of total variance; Cronbach's α=0.83; Table S15).

We also assessed the feasibility of applying the model for regulatory purposes by evaluating the performance of the model at three potential thresholds (Table S18). For a threshold value of 0.50, the model had a sensitivity of 81.5%, a specificity of 60.1%, a Positive Predictive Value (PPV) of 83.6%, and a Negative Predictive Value (NPV) of 56.5%, thus identifying 3,661 trials (69.6% of the hold out set). Decision curve analysis showed a net benefit for the model over the 'Flag All' and 'Flag None' scenarios across the clinically relevant range of probability thresholds (pt=0.10-0.55), therefore, suggesting that it has potential as a pre-registration time screening tool (Fig S8).

### SI.5. Economic Burden Cost Inputs and Assumptions

To convert all unreported studies to an identifiable measure of systematic waste through a quantifiable metric of wasted dollars per patient, estimates of costs for each study participant were developed from existing literature reference points based on a previously established benchmark value. Each estimate was developed as a function of the clinical phase, to reflect the different cost structures involved in Phase 1 vs. Phase 3 studies involving humans as participants. In order to calculate the total economic burden associated with each of the silent studies, the number of registered participants for each study was multiplied by the appropriate benchmark cost of each study participant as a function of the phase in which the study was conducted. An implicit assumption of the methodology is that the number of enrolled participants at the time of registration is very close to the final number of participants enrolled in the study. Furthermore, the calculations do not adjust for inflation as the benchmark values used are nominal and represent a base-line estimate only.

The remaining 200 studies whose clinical phases could not be determined were assumed to be classified as Phase 1 (lower bound), using the $20,000/participant as a conservative estimate. This and other assumptions regarding this issue are reported in detail in Table S25.

### SI.6. Sensitivity and Robustness Analyses

A second cohort for sensitivity testing was created to provide assurance of the robustness of the principal study results by limiting the analyses to those studies which had a 24 month gap from the Primary Completion Date to the end date for data extraction; this time period was used to confirm that studies which were classified as "silent" were abandoned and not simply experiencing typical delays in the review process for peer-reviewed publications. IPTW was then also applied to equally balance pre-existing covariates (including Phase, Regulatory Era, and Therapeutic Area), based on Baseline characteristics, across the High-OCI and Low-OCI treatment groups to isolate the independent influence of Operational Complexity. E-values were finally used to determine the minimum strength of association required of an unmeasured confounding variable so that it could completely nullify the positive association identified between the OCI Exposure and Trial Dissemination Probability.

### SI.7. Proportional Hazards Diagnostics

Scaled Schoenfeld residuals indicated violations of the proportional hazard assumptions regarding OCI (p-value < 0.001; $\chi^2$ = 58.05) and the FDA Era indicator (p-value < .001; $\chi^2$ = 96.52). Violations indicate the effect size of both variables are time-dependent, which would explain the changes observed by the Kaplan-Meier curve as the dissemination gap decreases between the high and low-OCI groups throughout the longer follow-up periods. As such, we recommend to interpret the Cox Hazard Ratio as an average effect across all study time points. Therefore, we chose the Relative Mean Survival Time (RMST) as our primary measure for assessing the absolute difference in timing of dissemination between groups.

### SI.8. Analytic Cohort Derivation and Missing Data Handling

The final analytic cohort was derived by sequentially applying the prespecified eligibility criteria to the operational dataset used in the analysis. A total of 34,741 study records were initially loaded. Restriction to completed trials yielded 23,759 records. Exclusion of records with missing primary completion dates left 23,551 trials. Restriction to trials completed on or before the analytic censor date left 22,779 trials. The cohort was then limited to studies belonging to the prespecified harmonized target sponsors, resulting in a final analytic sample of 17,537 trials.

These restrictions were applied to ensure that all included trials had a valid temporal anchor for dissemination assessment and belonged to the sponsor population defined a priori in the study design.

Missing data were handled according to the requirements of each analytic component while preserving the original prespecified pipeline. Trials missing the primary completion date were excluded from analyses requiring dissemination timing classification because temporal outcome assignment was not possible without this field. Phase values that could not be harmonized to a standard category were retained as an UNKNOWN category rather than excluded, in order to preserve analytic sample size. For derived structural variables contributing to the Operational Complexity Index (OCI), including enrollment, number of sites, and number of countries, the original operational transformation rules of the pipeline were retained unchanged for reproducibility.

The extent of missingness for variables used in the analytic workflow is reported in the supplementary missingness report. No matching procedures were used in this retrospective cohort design; therefore, STROBE item 6b was not applicable.

**SI.9. RECORD / STROBE Checklist Aligned to Manuscript**

| Section | Item No. | STROBE / RECORD item | Filled location in manuscript |
|---|---|---|---|
| **Title and Abstract** | 1a | Indicate study design in title or abstract | Title (“a retrospective cohort study”); Abstract, Methods and Findings (“We conducted a retrospective cohort study…”) |
| **Title and Abstract** | 1b | Provide informative summary in abstract | Abstract, Background; Methods and Findings; Conclusions |
| **Title and Abstract** | RECORD 1.1 | Type of data used and database names | Abstract, Methods and Findings: ClinicalTrials.gov registry data; manuscript specifies AACT in Methods – Data Source |
| **Title and Abstract** | RECORD 1.2 | Geographic region and timeframe | Abstract, Methods and Findings: “2007–2024”; “20 largest global pharmaceutical companies” |
| **Title and Abstract** | RECORD 1.3 | Data linkage statement | Not explicitly stated in the abstract; data linkage is described in Methods – Outcome Measures (ClinicalTrials.gov/AACT linked to PubMed using NCT identifiers) |
| **Introduction** | 2 | Scientific background and rationale | Introduction |
| **Introduction** | 3 | Objectives and hypotheses | End of Introduction |

| | | | |
|---|---|---|---|
| **Methods** | 4 | Study design presented early | Methods – Study Design and Reporting |
| **Methods** | 5 | Setting, location, relevant dates | Methods – Data Source; Methods – Cohort Selection |
| **Participants** | 6a | Eligibility criteria and participant selection | Methods – Cohort Selection; supplementary details in SI.8 |
| **Participants** | 6b | Matching criteria | Not applicable; no matching was used in this retrospective cohort design (SI.8) |
| **Participants** | RECORD 6.1 | Methods of study population selection | Methods – Cohort Selection; supplementary details in SI.1 and SI.8 |
| **Participants** | RECORD 6.2 | Validation of codes/algorithms | Methods – Outcome Measures; details in SI.2 |
| **Participants** | RECORD 6.3 | Data linkage flow diagram | A dedicated linkage flow diagram is not provided; Fig 1A presents cohort derivation rather than a linkage-specific flow diagram |
| **Variables** | 7 | Define outcomes, exposures, predictors, confounders | Methods – Exposure: OCI; Outcome Measures; Covariates |
| **Variables** | RECORD 7.1 | Codes/algorithms used to classify exposures/outcomes/confounders | Reported in part in Methods – Exposure, Outcome Measures, and Covariates; a fully enumerated code list is not provided in the main text |
| **Data Sources / Measurement** | 8 | Sources of data and assessment methods | Methods – Data Source; Outcome Measures |
| **Bias** | 9 | Efforts to address bias | Methods – Outcome Measures (manual |

| | | | |
|---|---|---|---|
| | | | validation); Methods – Sensitivity and Robustness Analyses (probabilistic bias analysis); Discussion – Limitations |
| **Study Size** | 10 | Explain study size | Methods – Cohort Selection; supplementary narrative cohort derivation in SI.8 |
| **Quantitative Variables** | 11 | Handling of quantitative variables | Methods – Exposure: OCI (log[x+1] transformation, standardization, PCA); Methods – Statistical Analysis |
| **Statistical Methods** | 12a | Statistical methods, including confounder control | Methods – Statistical Analysis |
| **Statistical Methods** | 12b | Methods for subgroups and interactions | Methods – Sensitivity and Robustness Analyses (phase-stratified analyses; oncology-only analyses) |
| **Statistical Methods** | 12c | Explain how missing data were addressed | Supplementary Methods – SI.8; Supplementary Missingness Report |
| **Statistical Methods** | 12d | Loss to follow-up | Not applicable for this registry-based retrospective cohort of completed trials |
| **Statistical Methods** | 12e | Sensitivity analyses | Methods – Sensitivity and Robustness Analyses |
| **Statistical Methods** | RECORD 12.1 | Extent of investigators' access to database population | Methods – Data Source indicates use of a static AACT data extract; a separate formal access statement is not provided |
| **Statistical Methods** | RECORD 12.2 | Data cleaning methods | Methods – Cohort Selection (sponsor harmonization, exclusions); Methods – Data |

| | | | |
|---|---|---|---|
| | | | Source; supplementary clarification in SI.8 |
| **Statistical Methods** | RECORD 12.3 | Linkage methods and quality evaluation | Methods – Outcome Measures (PubMed linkage using NCT identifiers); linkage evaluation and validation described in the same section and in SI.2 |
| **Results** | 13a | Report numbers of individuals at each stage | Results – Baseline Characteristics and Automated Pipeline Validation; Fig 1A; supplementary narrative cohort derivation in SI.8 |
| **Results** | 13b | Give reasons for non-participation/non-inclusion | Methods – Cohort Selection; Fig 1A; supplementary narrative exclusions in SI.8 |
| **Results** | 13c | Consider use of a flow diagram | Fig 1A |
| **Results** | RECORD 13.1 | Detailed selection of persons included | Fig 1A; Methods – Cohort Selection; supplementary details in SI.1 and SI.8 |
| **Descriptive Data** | 14a | Characteristics of study participants / trials | Results – Baseline Characteristics and Automated Pipeline Validation; Table 1 |
| **Descriptive Data** | 14b | Number with missing data for each variable | Supplementary Missingness Report; Supplementary Methods – SI.8 |
| **Descriptive Data** | 14c | Follow-up time | Methods – Outcome Measures defines time to dissemination and censoring; Results – Temporal Dynamics and Time-to-Dissemination |

| | | | |
|---|---|---|---|
| **Outcome Data** | 15 | Report outcome events / summary measures over time | Results – Baseline Characteristics and Automated Pipeline Validation; Results – Temporal Dynamics and Time-to-Dissemination; Table 1; Table 2; Figs 1–3 |
| **Main Results** | 16a | Unadjusted/adjusted estimates with precision | Results – Operational Complexity and Dissemination Probability; Results – Temporal Dynamics and Time-to-Dissemination; Table 2 |
| **Main Results** | 16b | Report category boundaries when continuous variables categorized | Results – OCI deciles and tertiles in Figs 2 and 3 |
| **Main Results** | 16c | Translate relative risk into absolute risk where relevant | Results – Temporal Dynamics and Time-to-Dissemination via RMST difference |
| **Other Analyses** | 17 | Subgroup analyses and sensitivity analyses | Results – Predictive Modeling; Results – Economic Burden of Non-Dissemination; Results – Robustness and Sensitivity Analyses |
| **Discussion** | 18 | Summarize key results with reference to objectives | Discussion – Summary of Findings |
| **Discussion** | 19 | Discuss limitations | Discussion – Limitations |
| **Discussion** | RECORD 19.1 | Discuss implications of using routinely collected data | Discussion – Limitations; also addressed in Methods – Data Source and Outcome Measures |
| **Discussion** | 20 | Overall interpretation | Discussion; Conclusion |

| | | | |
|---|---|---|---|
| **Discussion** | 21 | Generalisability | Discussion – Limitations (restricted to the 20 largest pharmaceutical sponsors) |
| **Other Information** | 22 | Funding | Funding statement at end of manuscript (“This study received no external funding…”) |
| **Other Information** | RECORD 22.1 | Access to protocol, raw data, or programming code | Methods – final paragraph (“master_pipeline.py … supplementary repository”) |

### Supplementary Figs

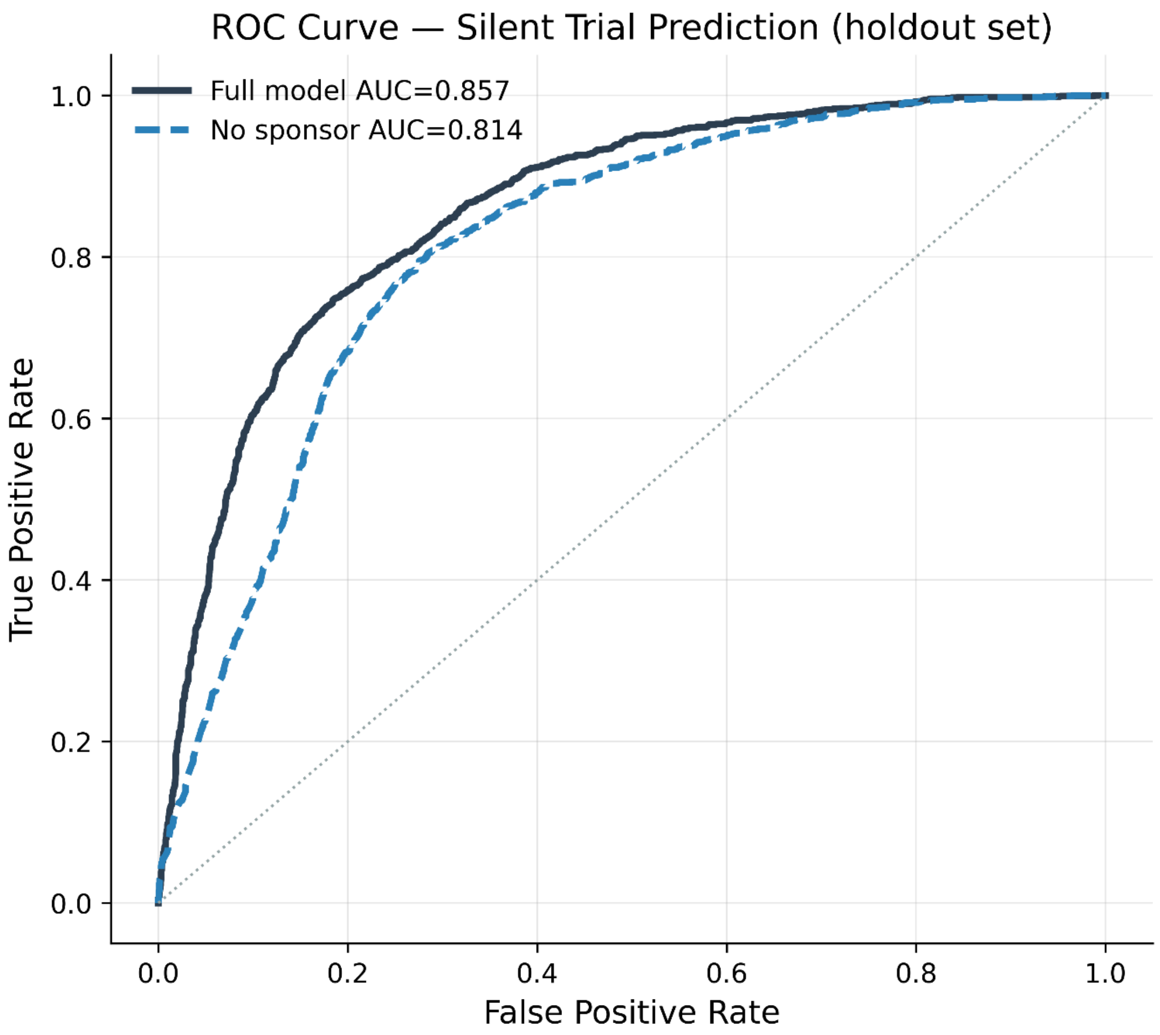


**Fig S1. Receiver Operating Characteristic Curves for Prediction of Silent Trial Dissemination**

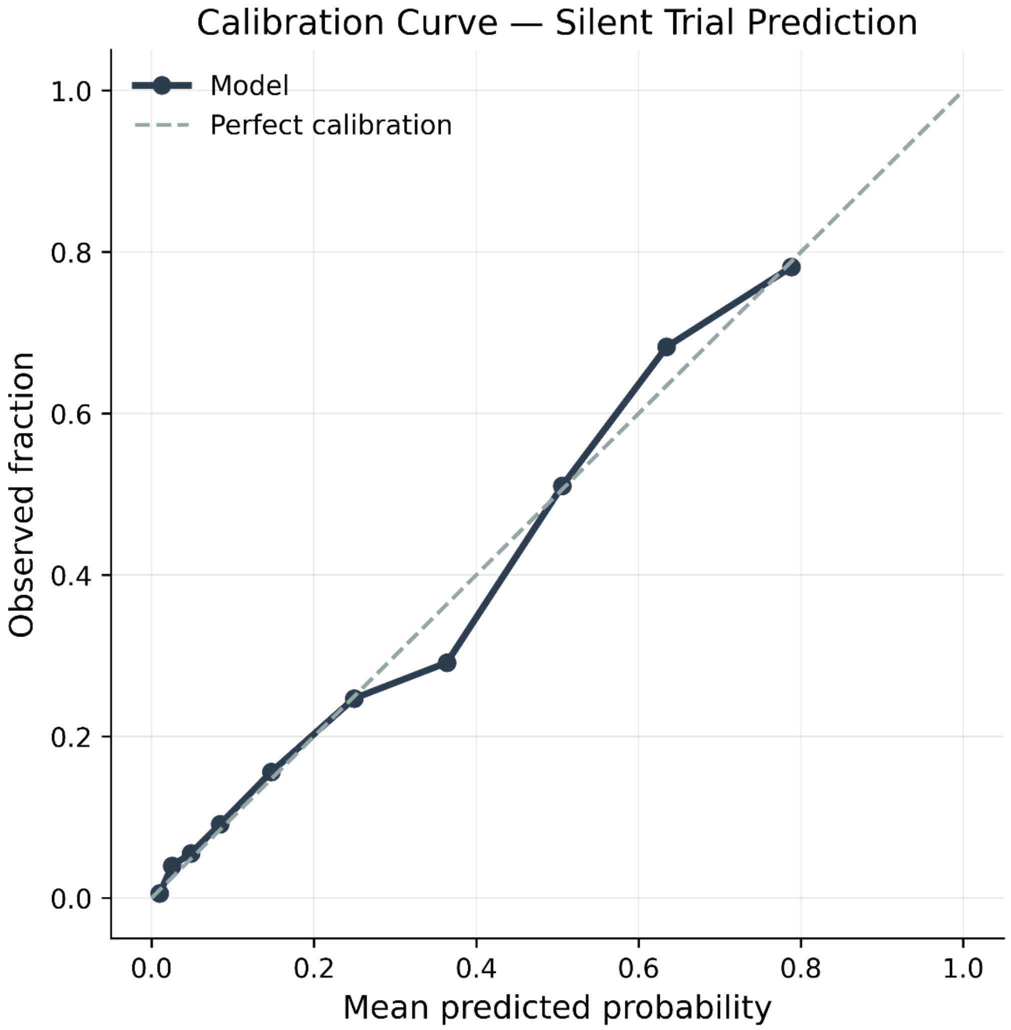


**Fig S2. Calibration Curve for the Predictive Model of Silent Trial Dissemination**

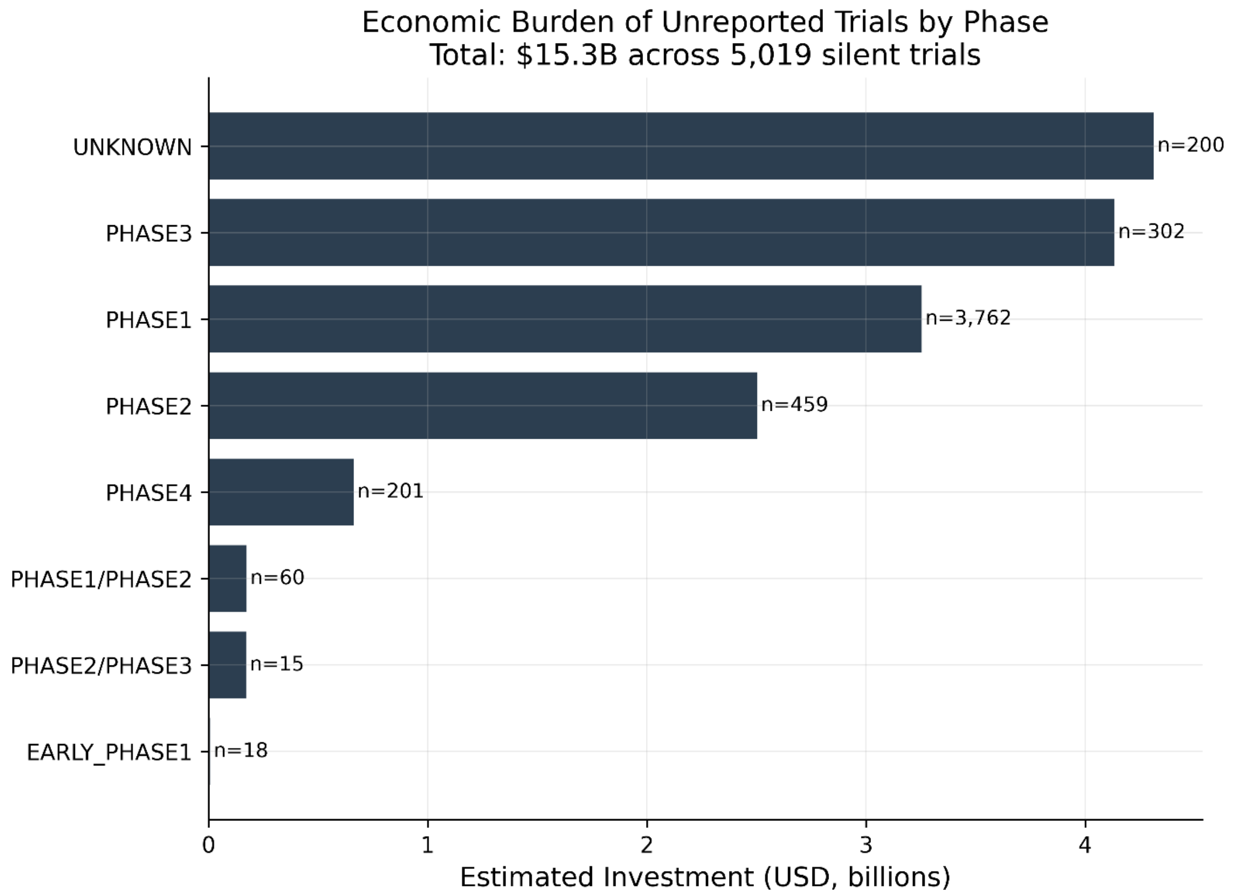


**Fig S3. Estimated Economic Burden of Silent Trials Stratified by Clinical Phase**

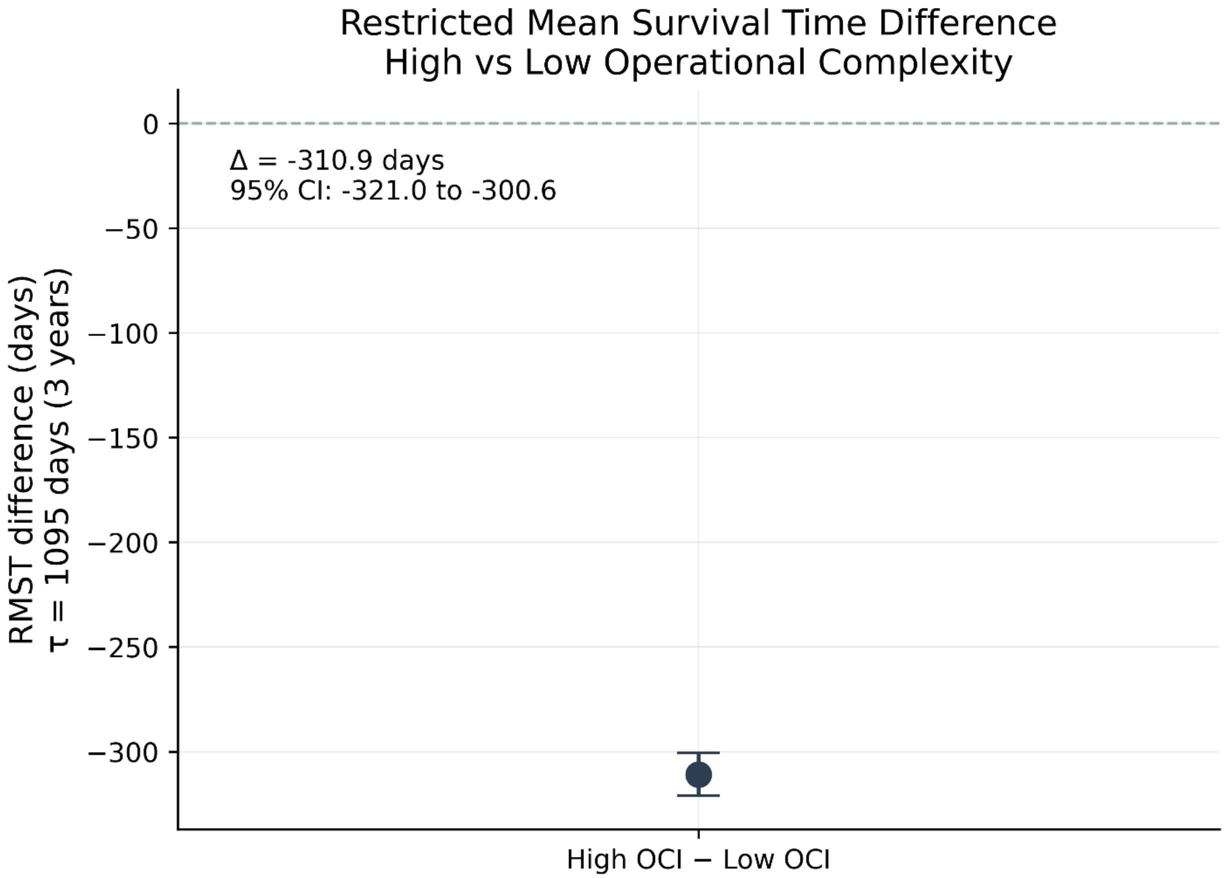


**Fig S4. Restricted Mean Survival Time Difference Between High- and Low-OCI Trials**

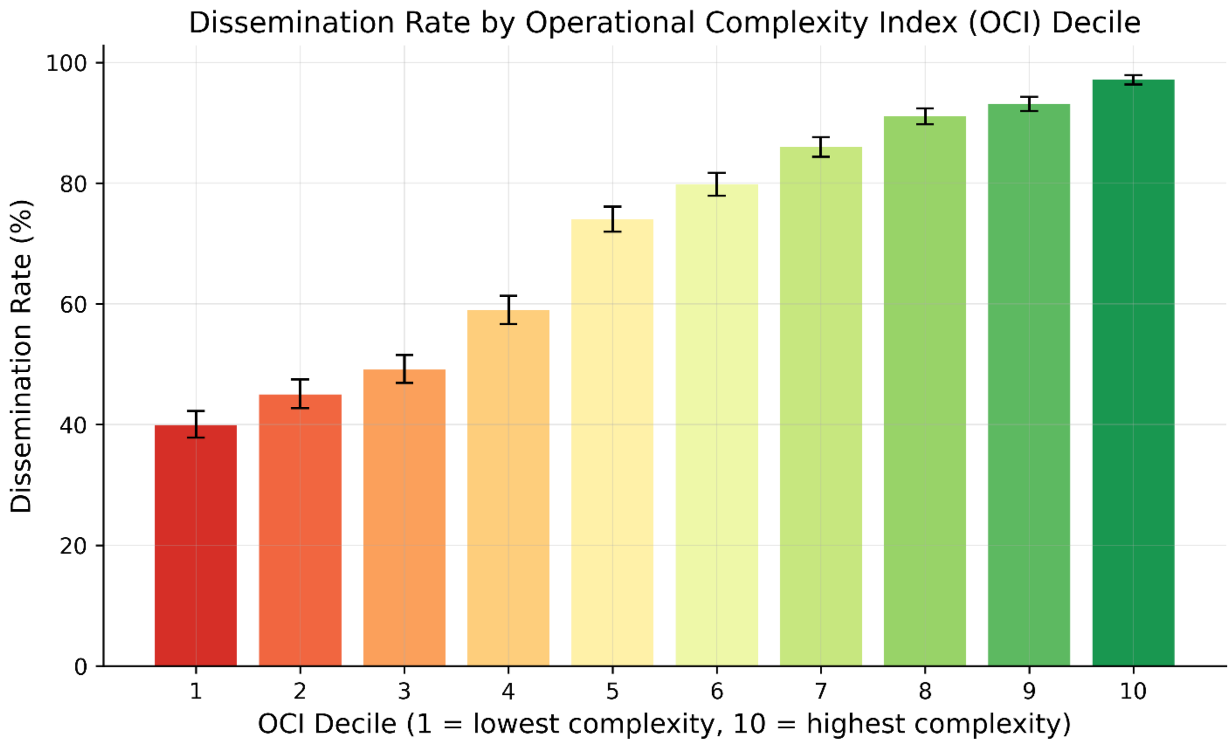


**Fig S5. Dissemination Rate by Operational Complexity Index Decile**

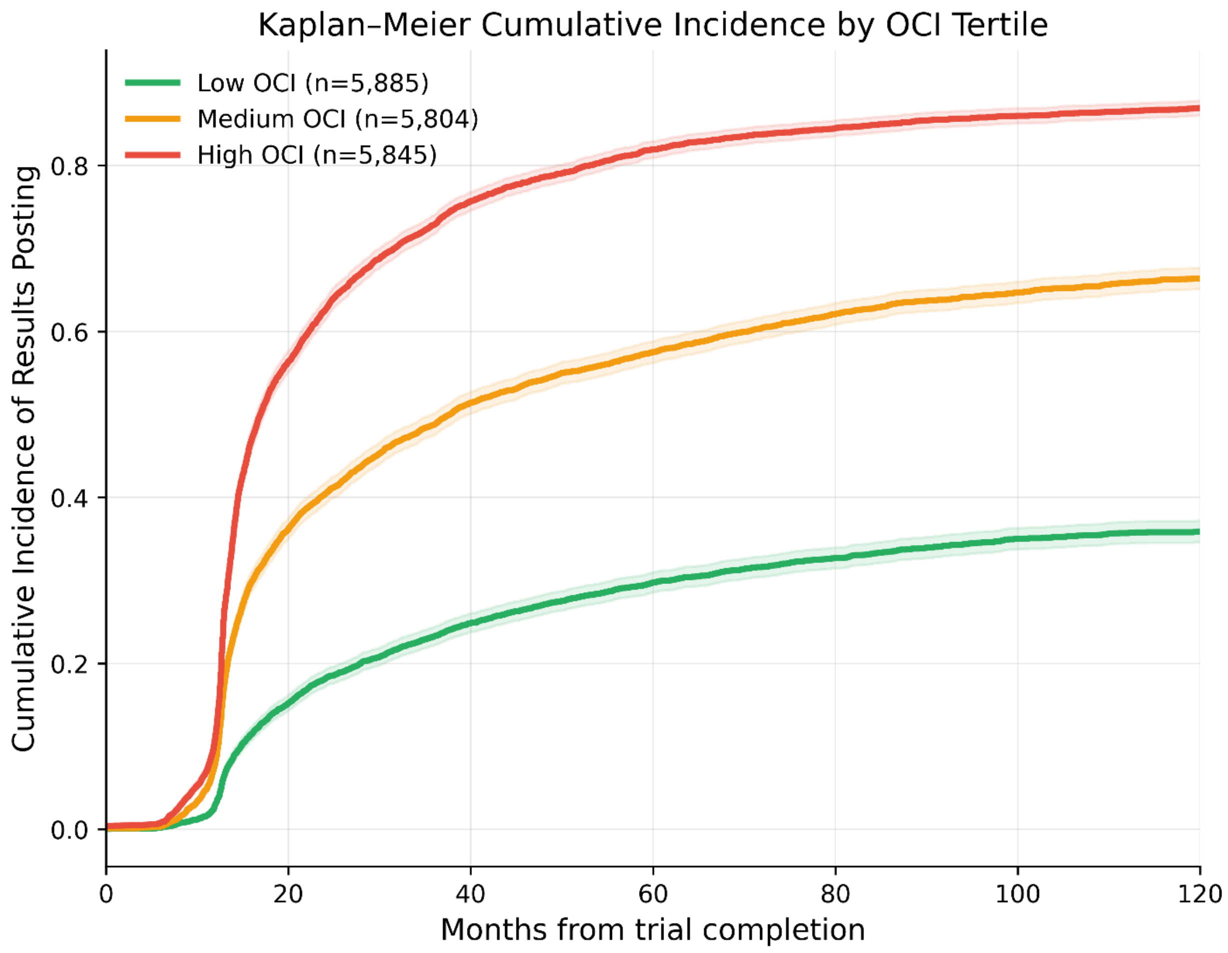


**Fig S6. Cumulative Incidence of Registry Results Posting by OCI Tertile**

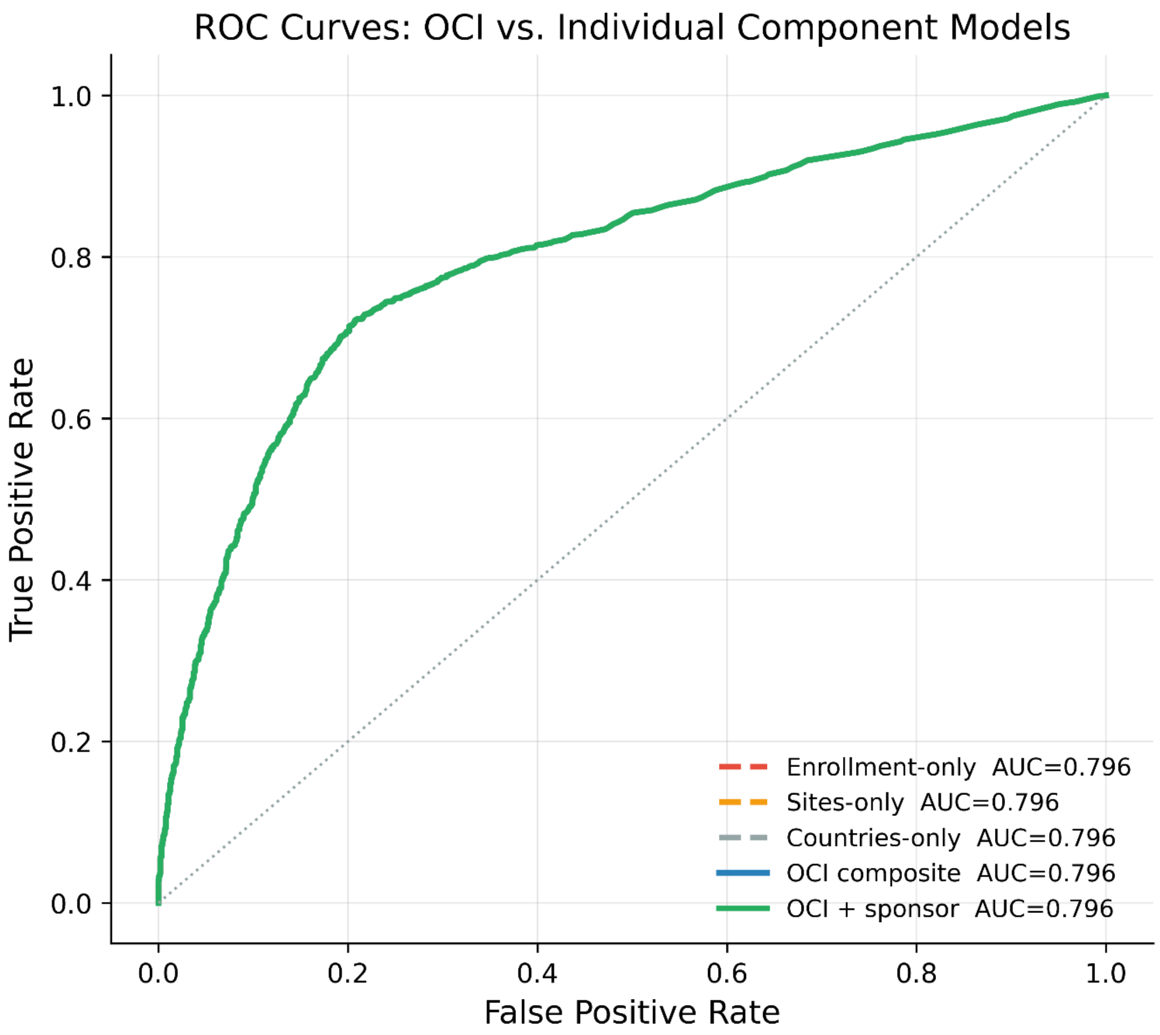


**Fig S7. Receiver Operating Characteristic Curves for Single-Component and Composite OCI Models**

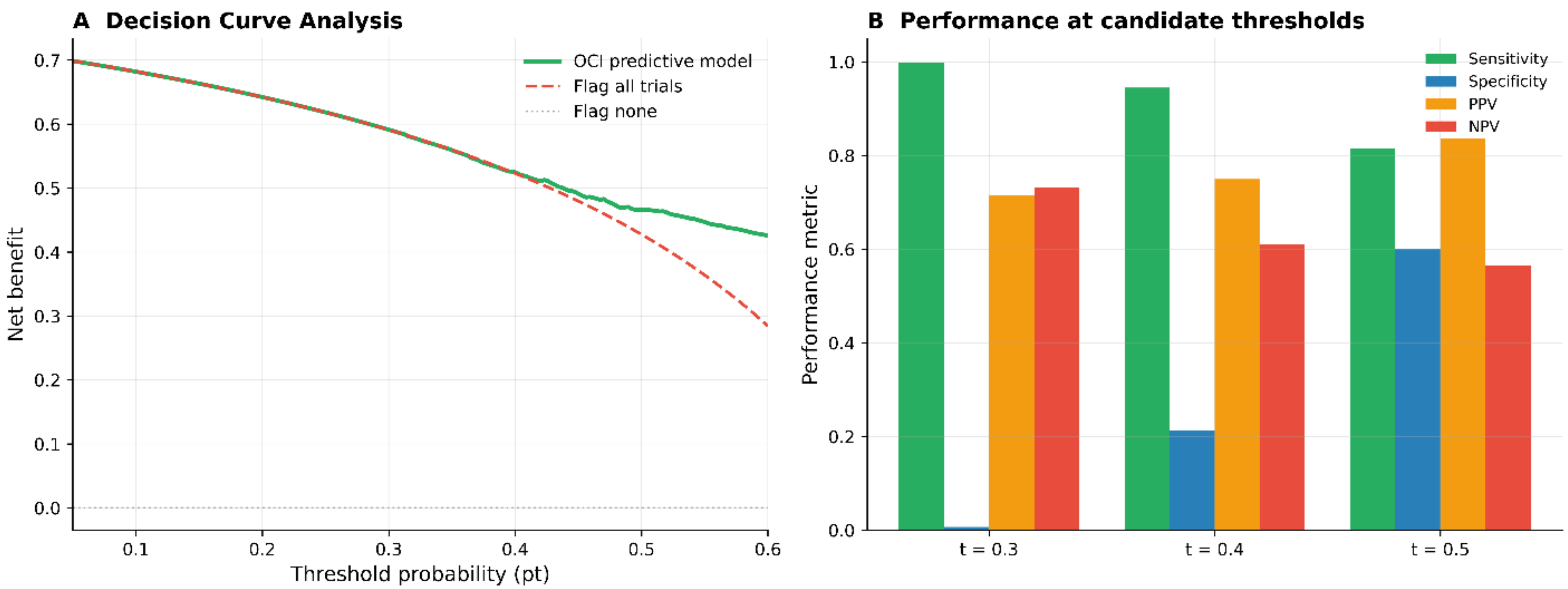


296

297 **Fig S8. Threshold Performance and Decision Curve Analysis**

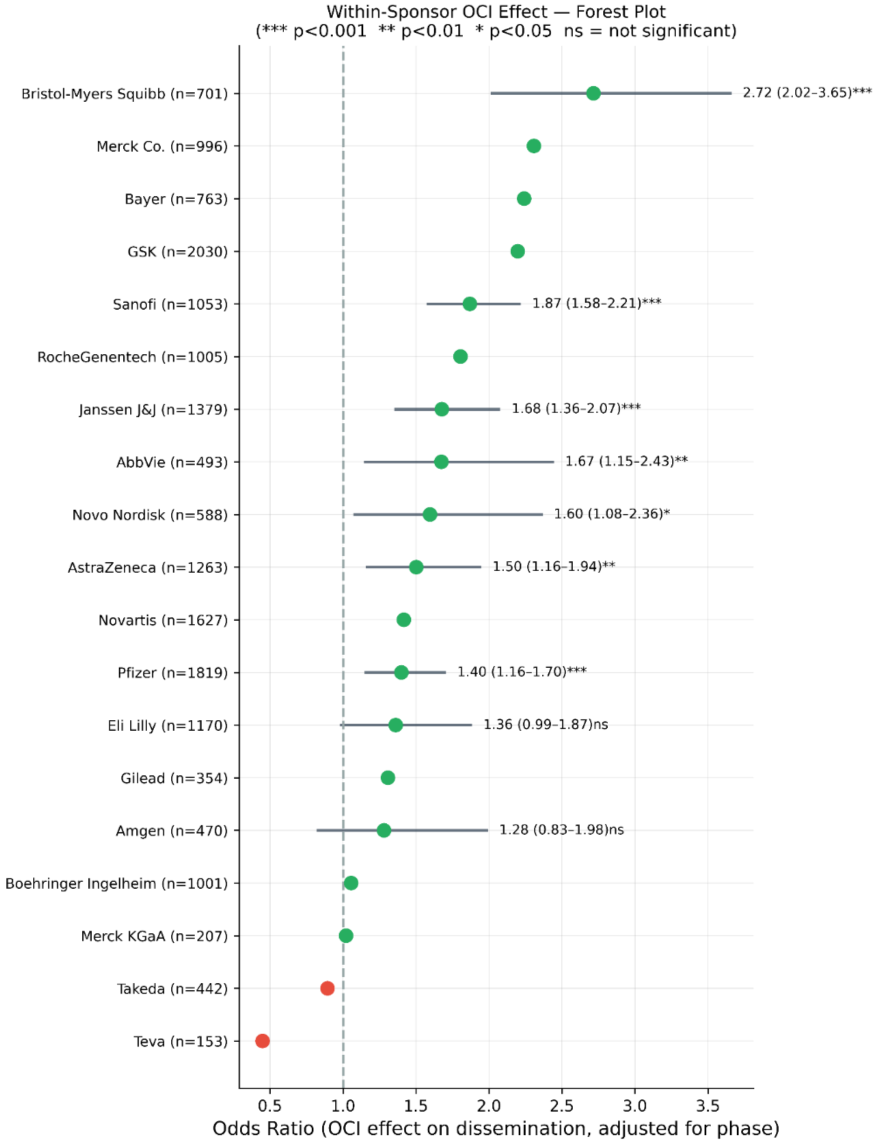


**Fig S9. Sponsor-Specific Adjusted Odds Ratios for Trial Dissemination**

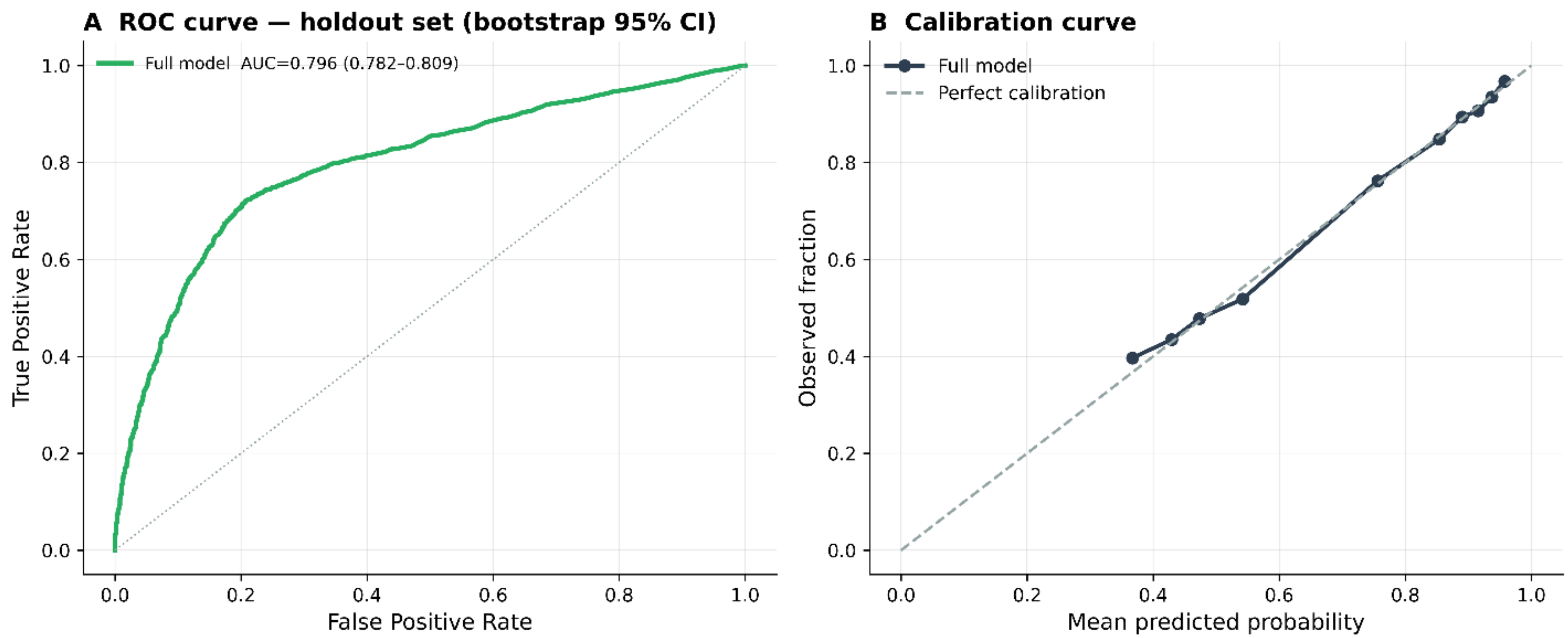


**Fig S10. Bootstrap Holdout AUC and Calibration Performance of the Predictive Model**

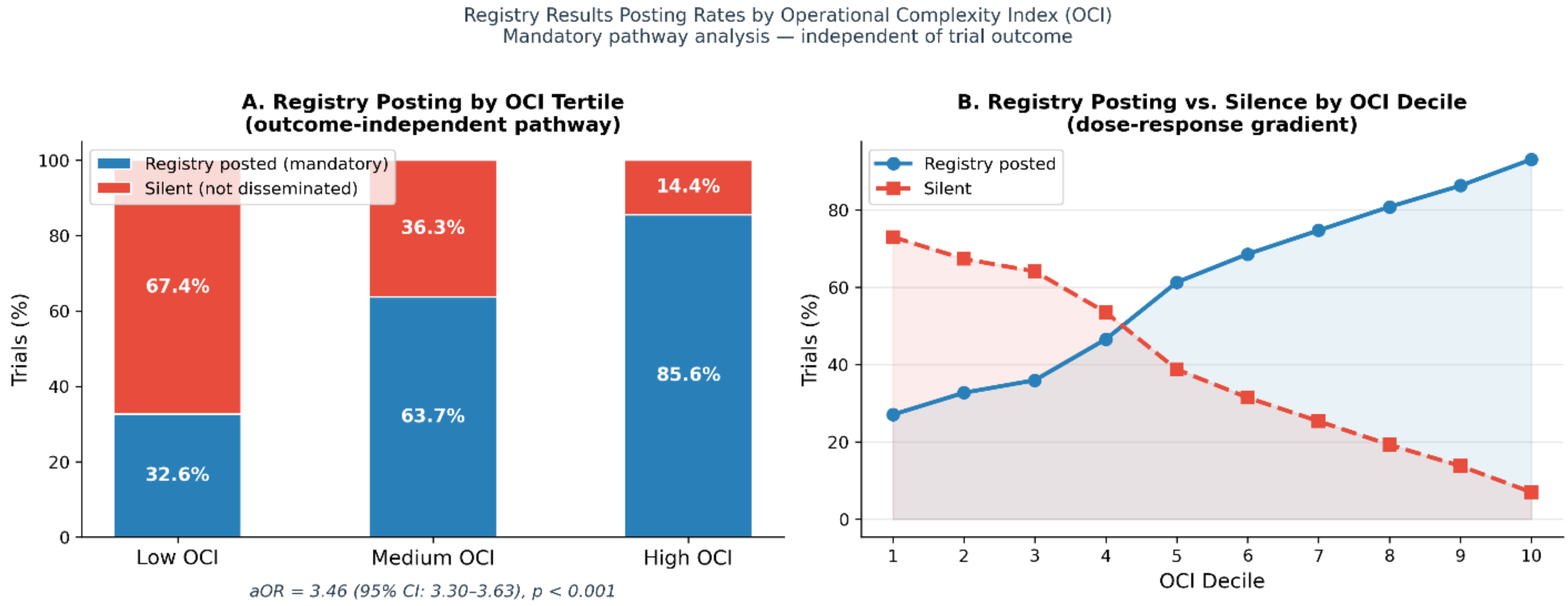


**Fig S11. Registry Results Posting Rates Stratified by Operational Complexity Index**

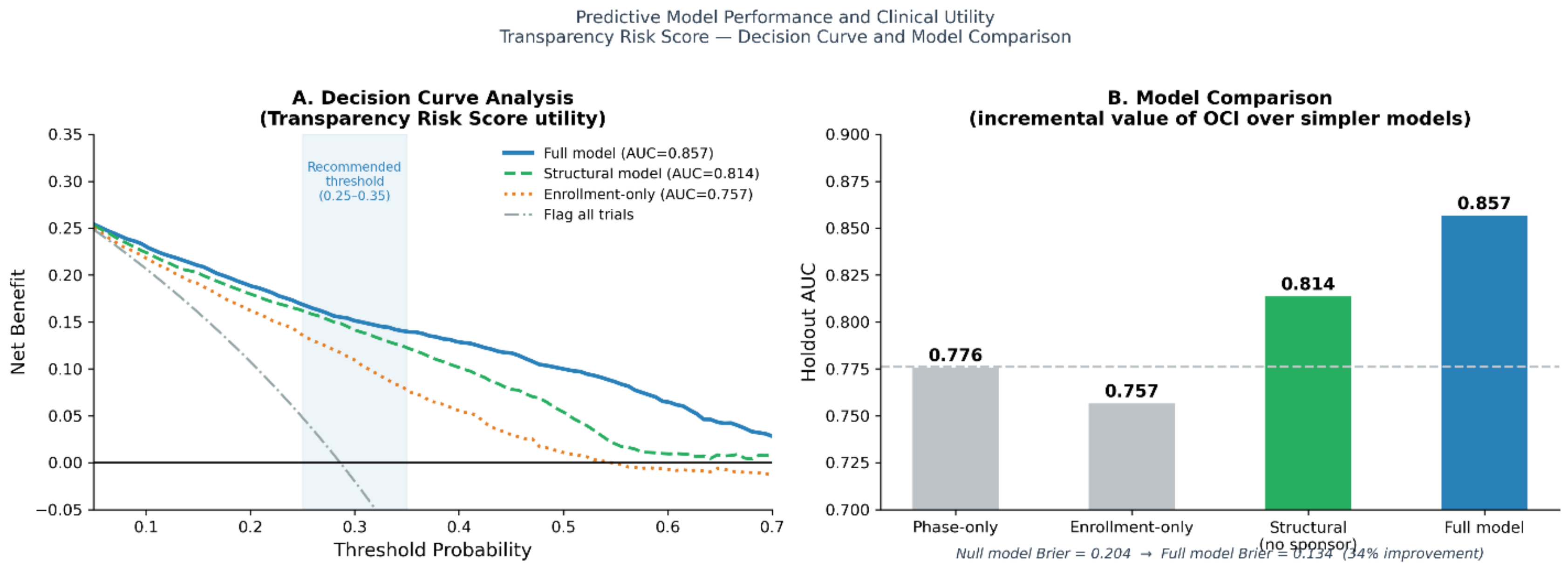


**Fig S12. Predictive Model Performance and Clinical Utility of the Transparency Risk Score**

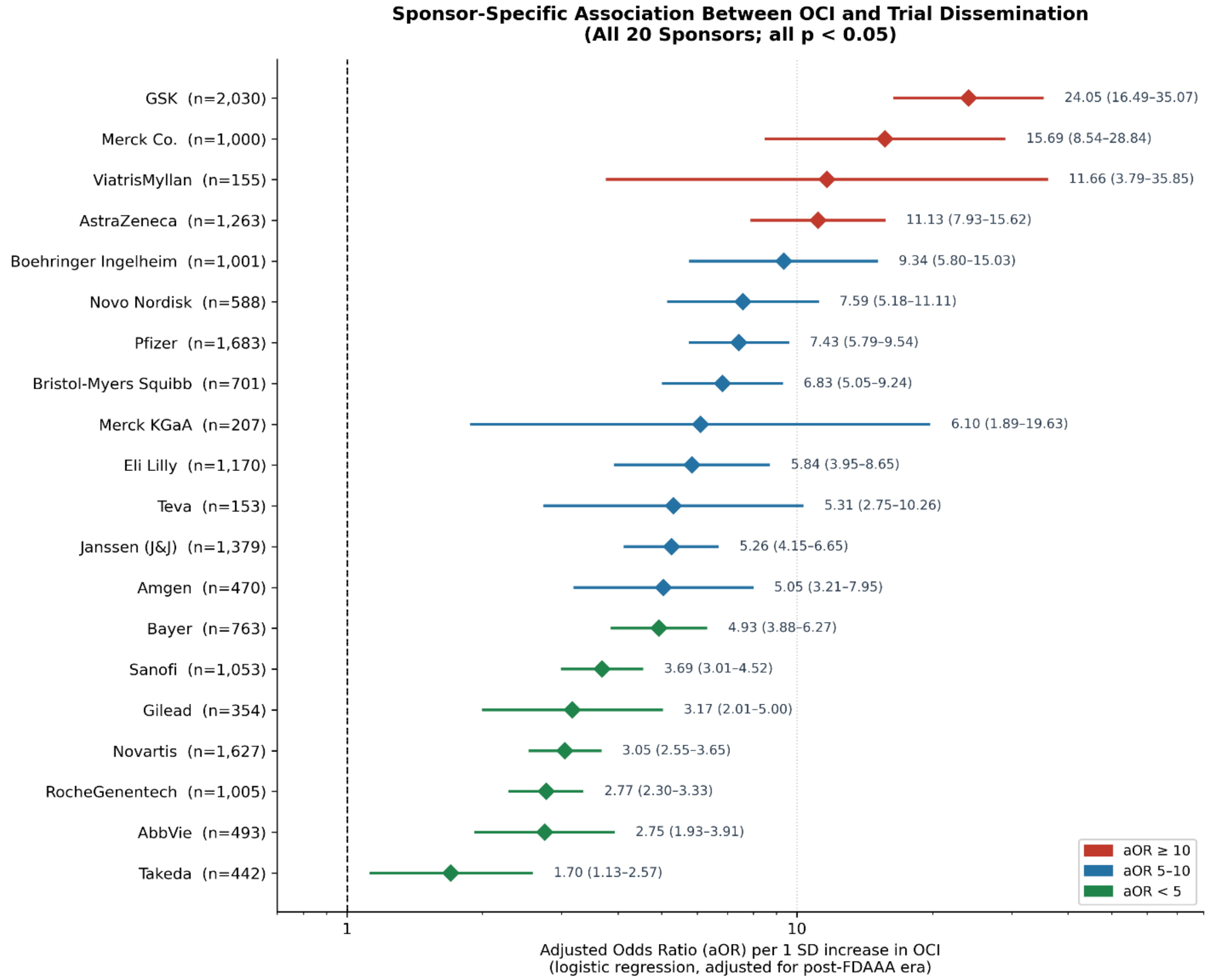


**Fig S13. Sponsor-Specific Association Between Operational Complexity and Trial Dissemination Across the 20 Largest Pharmaceutical Sponsors**

### Supplementary Tables

**Table S1. Baseline Characteristics of the Analytic Trial Cohort**

| Characteristic | N | Pct |
|---|---|---|
| **Total trials** | 17537 | |
| **Disseminated (any route)** | 12518 | 71.4% |
| **Silent (not disseminated)** | 5019 | 28.6% |
| **Registry results posted** | 10633 | 60.6% |
| **Post-FDAAA (start ≥ 2008)** | 14995 | 85.5% |
| **Phase: EARLY_PHASE1** | 29 | 0.2% |
| **Phase: PHASE1** | 6916 | 39.4% |
| **Phase: PHASE1/PHASE2** | 339 | 1.9% |
| **Phase: PHASE2** | 3430 | 19.6% |
| **Phase: PHASE2/PHASE3** | 129 | 0.7% |
| **Phase: PHASE3** | 4584 | 26.1% |
| **Phase: PHASE4** | 1529 | 8.7% |
| **Phase: UNKNOWN** | 581 | 3.3% |
| **OCI mean (SD)** | 0.00 (0.88) | |
| **Median enrollment (IQR)** | 79 (32–274) | |
| **Sponsor: AbbVie** | 493 | 2.8% |
| **Sponsor: Amgen** | 470 | 2.7% |
| **Sponsor: AstraZeneca** | 1263 | 7.2% |
| **Sponsor: Bayer** | 763 | 4.4% |
| **Sponsor: Bristol-Myers Squibb** | 701 | 4.0% |
| **Sponsor: Boehringer Ingelheim** | 1001 | 5.7% |
| **Sponsor: Eli Lilly** | 1170 | 6.7% |
| **Sponsor: Gilead** | 354 | 2.0% |
| **Sponsor: GSK** | 2030 | 11.6% |
| **Sponsor: Janssen (J&J)** | 1379 | 7.9% |
| **Sponsor: Merck & Co.** | 1000 | 5.7% |
| **Sponsor: Merck KGaA** | 207 | 1.2% |
| **Sponsor: Novartis** | 1627 | 9.3% |
| **Sponsor: Novo Nordisk** | 588 | 3.4% |
| **Sponsor: Pfizer** | 1683 | 9.6% |
| **Sponsor: Roche/Genentech** | 1005 | 5.7% |
| **Sponsor: Sanofi** | 1053 | 6.0% |
| **Sponsor: Takeda** | 442 | 2.5% |

| | | |
|---|---|---|
| **Sponsor: Teva** | 153 | 0.9% |
| **Sponsor: Viatris/Mylan** | 155 | 0.9% |

**Table S2. Annual Trial Initiation Volume by Sponsor, 2007–2024**

| sponsor_group | 2007 | 2008 | 2009 | 2010 | 2011 | 2012 | 2013 | 2014 | 2015 | 2016 | 2017 | 2018 | 2019 | 2020 | 2021 | 2022 | 2023 | 2024 | All |
|---|---|---|---|---|---|---|---|---|---|---|---|---|---|---|---|---|---|---|---|
| abbvie | 15 | 25 | 26 | 32 | 28 | 40 | 31 | 41 | 46 | 33 | 38 | 25 | 28 | 27 | 16 | 18 | 18 | 6 | 493 |
| amgen | 35 | 29 | 43 | 36 | 27 | 37 | 41 | 29 | 31 | 27 | 24 | 21 | 27 | 18 | 21 | 12 | 6 | 6 | 470 |
| astrazeneca | 73 | 163 | 119 | 142 | 89 | 59 | 62 | 88 | 80 | 42 | 53 | 37 | 50 | 58 | 59 | 46 | 22 | 21 | 1263 |
| bayer | 51 | 48 | 65 | 50 | 55 | 32 | 55 | 42 | 92 | 66 | 38 | 49 | 31 | 27 | 26 | 16 | 7 | 13 | 763 |
| bms | 43 | 50 | 50 | 40 | 51 | 48 | 32 | 60 | 37 | 50 | 56 | 38 | 41 | 28 | 18 | 24 | 30 | 5 | 701 |
| boehringer | 70 | 102 | 91 | 86 | 77 | 72 | 58 | 61 | 40 | 55 | 49 | 33 | 49 | 31 | 39 | 30 | 37 | 21 | 1001 |
| eli_lilly | 67 | 71 | 75 | 98 | 94 | 73 | 85 | 52 | 71 | 63 | 64 | 61 | 61 | 67 | 59 | 49 | 46 | 14 | 1170 |
| gilead | 9 | 23 | 7 | 20 | 38 | 36 | 40 | 38 | 55 | 34 | 18 | 11 | 10 | 8 | 4 | 1 | 2 | 0 | 354 |
| gsk | 245 | 216 | 246 | 212 | 181 | 152 | 150 | 132 | 97 | 109 | 83 | 48 | 44 | 22 | 30 | 29 | 22 | 12 | 2030 |
| janssen | 83 | 95 | 73 | 88 | 95 | 93 | 70 | 99 | 113 | 85 | 95 | 82 | 67 | 68 | 94 | 47 | 26 | 6 | 1379 |
| merck | 121 | 131 | 87 | 74 | 68 | 62 | 54 | 55 | 46 | 39 | 28 | 43 | 48 | 33 | 22 | 32 | 34 | 23 | 1000 |
| merck_kgaa | 14 | 21 | 20 | 7 | 12 | 9 | 15 | 14 | 13 | 5 | 5 | 22 | 12 | 10 | 12 | 11 | 4 | 1 | 207 |
| novartis | 145 | 169 | 169 | 165 | 146 | 164 | 134 | 102 | 96 | 70 | 70 | 52 | 45 | 29 | 41 | 18 | 10 | 2 | 1627 |
| novo_nordisk | 28 | 29 | 46 | 43 | 33 | 44 | 47 | 40 | 23 | 40 | 32 | 29 | 38 | 22 | 38 | 23 | 25 | 8 | 588 |
| pfizer | 154 | 189 | 191 | 160 | 148 | 104 | 98 | 91 | 65 | 49 | 59 | 61 | 62 | 71 | 65 | 49 | 47 | 20 | 1683 |
| roche | 79 | 98 | 80 | 77 | 117 | 92 | 83 | 73 | 56 | 59 | 45 | 39 | 34 | 36 | 19 | 14 | 3 | 1 | 1005 |
| sanofi | 110 | 116 | 102 | 100 | 72 | 84 | 54 | 47 | 66 | 50 | 37 | 49 | 41 | 44 | 40 | 22 | 14 | 5 | 1053 |
| takeda | 33 | 24 | 22 | 20 | 31 | 26 | 15 | 41 | 49 | 34 | 30 | 31 | 13 | 14 | 26 | 14 | 16 | 3 | 442 |
| teva | 15 | 7 | 12 | 17 | 8 | 12 | 26 | 14 | 10 | 7 | 4 | 3 | 4 | 4 | 3 | 4 | 1 | 2 | 153 |
| viatris | 20 | 29 | 23 | 21 | 23 | 10 | 4 | 1 | 2 | 4 | 0 | 6 | 3 | 3 | 5 | 0 | 1 | 0 | 155 |
| All | 1410 | 1635 | 1547 | 1488 | 1393 | 1249 | 1154 | 1120 | 1088 | 921 | 828 | 740 | 708 | 620 | 637 | 459 | 371 | 169 | 17537 |

**Table S3. Correlation Matrix for Operational Complexity Index Components**

| Variable | log enrollment | log sites | log countries | OCI (standardized) |
|---|---|---|---|---|
| **log enrollment** | **1.000** | **0.660** | **0.515** | **0.825** |
| **log sites** | **0.660** | **1.000** | **0.799** | **0.933** |
| **log countries** | **0.515** | **0.799** | **1.000** | **0.878** |
| **OCI (standardized)** | **0.825** | **0.933** | **0.878** | **1.000** |

**Table S4. Multivariable Logistic Regression Model for Trial Dissemination**

| Term | Coefficient | Odds ratio | 95% CI | p-value |
|---|---|---|---|---|
| **Intercept** | -0.693 | 0.500 | 0.203–1.236 | 0.133 |
| **Operational Complexity Index (standardized)** | 0.883 | 2.419 | 2.238–2.613 | <0.001 |
| **Post-FDAAA era** | 0.695 | 2.004 | 1.785–2.250 | <0.001 |
| **Phase 1** | 0.339 | 1.404 | 0.610–3.230 | 0.425 |
| **Phase 1/2** | 1.864 | 6.450 | 2.653–15.679 | <0.001 |
| **Phase 2** | 2.032 | 7.632 | 3.296–17.673 | <0.001 |
| **Phase 2/3** | 1.843 | 6.313 | 2.278–17.495 | <0.001 |
| **Phase 3** | 2.604 | 13.514 | 5.810–31.435 | <0.001 |
| **Phase 4** | 2.498 | 12.159 | 5.206–28.400 | <0.001 |
| **Unknown phase** | 1.849 | 6.355 | 2.706–14.925 | <0.001 |
| **Sponsor: Amgen** | 0.458 | 1.581 | 1.079–2.317 | 0.019 |
| **Sponsor: AstraZeneca** | 0.218 | 1.243 | 0.918–1.683 | 0.159 |
| **Sponsor: Bayer** | -1.660 | 0.190 | 0.138–0.262 | <0.001 |
| **Sponsor: Bristol-Myers Squibb** | -0.786 | 0.456 | 0.327–0.634 | <0.001 |
| **Sponsor: Boehringer Ingelheim** | 1.145 | 3.143 | 2.295–4.304 | <0.001 |
| **Sponsor: Eli Lilly** | 1.421 | 4.143 | 3.015–5.693 | <0.001 |
| **Sponsor: Gilead** | 0.198 | 1.219 | 0.790–1.881 | 0.370 |
| **Sponsor: GSK** | 0.313 | 1.367 | 1.020–1.833 | 0.037 |
| **Sponsor: Janssen (Johnson & Johnson)** | -0.400 | 0.670 | 0.496–0.905 | 0.009 |

| | | | | |
|---|---|---|---|---|
| **Sponsor: Merck & Co.** | 1.232 | 3.429 | 2.471–4.756 | <0.001 |
| **Sponsor: Merck KGaA** | 1.648 | 5.199 | 3.061–8.828 | <0.001 |
| **Sponsor: Novartis** | -0.674 | 0.510 | 0.379–0.686 | <0.001 |
| **Sponsor: Novo Nordisk** | -0.251 | 0.778 | 0.545–1.111 | 0.167 |
| **Sponsor: Pfizer** | 0.270 | 1.310 | 0.979–1.755 | 0.070 |
| **Sponsor: Roche/Genentech** | -0.544 | 0.580 | 0.424–0.794 | <0.001 |
| **Sponsor: Sanofi** | -1.356 | 0.258 | 0.190–0.350 | <0.001 |
| **Sponsor: Takeda** | 1.303 | 3.680 | 2.430–5.573 | <0.001 |
| **Sponsor: Teva** | -0.069 | 0.933 | 0.526–1.656 | 0.813 |
| **Sponsor: Viatris/Mylan** | 0.294 | 1.341 | 0.814–2.211 | 0.250 |
| **Therapeutic area: Immunology** | 0.194 | 1.215 | 0.915–1.613 | 0.179 |
| **Therapeutic area: Infectious disease** | 0.370 | 1.447 | 1.140–1.838 | 0.002 |
| **Therapeutic area: Metabolic** | -0.121 | 0.886 | 0.699–1.123 | 0.317 |
| **Therapeutic area: Neurology** | -0.154 | 0.857 | 0.668–1.100 | 0.227 |
| **Therapeutic area: Oncology** | 0.452 | 1.572 | 1.253–1.972 | <0.001 |
| **Therapeutic area: Other** | -0.217 | 0.805 | 0.654–0.990 | 0.040 |
| **Therapeutic area: Respiratory** | 0.073 | 1.076 | 0.824–1.405 | 0.590 |

**Table S5. Cox Proportional Hazards Model for Time to Dissemination**

| Covariate | Hazard Ratio (HR) | 95% CI | p-value |
|---|---|---|---|
| **Operational Complexity (OCI, per 1 SD)** | **1.25** | 1.22–1.28 | <0.001 |
| **Post-FDAAA era** | 1.83 | 1.73–1.94 | <0.001 |
| **Phase 1** | 1.38 | 0.57–3.32 | 0.475 |
| **Phase 1/2** | 5.48 | 2.26–13.31 | <0.001 |
| **Phase 2** | 5.09 | 2.12–12.26 | <0.001 |

| | | | |
|---|---|---|---|
| **Phase 2/3** | 6.39 | 2.60–15.69 | <0.001 |
| **Phase 3** | 7.77 | 3.23–18.71 | <0.001 |
| **Phase 4** | 9.30 | 3.86–22.41 | <0.001 |
| **Phase unknown** | 6.74 | 2.79–16.31 | <0.001 |
| **Amgen** | 1.22 | 1.04–1.42 | 0.014 |
| **AstraZeneca** | 1.07 | 0.93–1.23 | 0.346 |
| **Bayer** | 0.28 | 0.24–0.34 | <0.001 |
| **Bristol Myers Squibb** | 0.71 | 0.61–0.83 | <0.001 |
| **Boehringer Ingelheim** | 1.77 | 1.55–2.03 | <0.001 |
| **Eli Lilly** | 1.75 | 1.54–2.00 | <0.001 |
| **Gilead** | 1.34 | 1.14–1.58 | <0.001 |
| **GSK** | 1.03 | 0.91–1.17 | 0.655 |
| **Janssen** | 0.67 | 0.58–0.77 | <0.001 |
| **Merck** | 1.97 | 1.72–2.26 | <0.001 |
| **Merck KGaA** | 1.88 | 1.56–2.27 | <0.001 |
| **Novartis** | 0.79 | 0.70–0.90 | <0.001 |
| **Novo Nordisk** | 0.76 | 0.64–0.90 | 0.002 |
| **Pfizer** | 1.16 | 1.02–1.32 | 0.026 |
| **Roche / Genentech** | 0.63 | 0.55–0.73 | <0.001 |
| **Sanofi** | 0.37 | 0.32–0.43 | <0.001 |
| **Takeda** | 1.43 | 1.22–1.66 | <0.001 |
| **Teva** | 0.78 | 0.63–0.97 | 0.024 |
| **Viatris** | 1.35 | 1.07–1.71 | 0.011 |
| **Immunology** | 1.05 | 0.94–1.18 | 0.401 |
| **Infectious disease** | 1.10 | 0.99–1.21 | 0.064 |
| **Metabolic** | 0.84 | 0.76–0.93 | <0.001 |

| | | | |
|---|---|---|---|
| **Neurology** | 0.87 | 0.78–0.97 | 0.010 |
| **Oncology** | 1.13 | 1.02–1.24 | 0.016 |
| **Other** | 1.00 | 0.91–1.10 | 0.986 |
| **Respiratory** | 0.98 | 0.88–1.09 | 0.719 |

Cox proportional hazards model evaluating factors associated with time to dissemination of trial results. Hazard ratios represent relative differences in dissemination timing. Values greater than 1 indicate earlier dissemination compared with the reference category.

**Table S6. Restricted Mean Survival Time Analysis Comparing High- and Low-OCI Trials**

| Time horizon, days | RMST, low OCI (days) | RMST, high OCI (days) | RMST difference, days (high − low) | 95% CI | Total trials, n | Low OCI trials, n | High OCI trials, n |
|---|---|---|---|---|---|---|---|
| **1,095** | 969.983 | 659.103 | -310.880 | -320.959 to -300.588 | 17,534 | 5,885 | 5,845 |

**Table S7. Predictive Performance of Machine Learning Models in Cross-Validation and Holdout Testing**

| Model | CV AUC (mean) | CV AUC (SD) | Holdout AUC | Brier Score | N |
|---|---|---|---|---|---|
| **Full (all features incl. sponsor)** | 0.858 | 0.005 | 0.857 | 0.134 | 17,534 |
| **Structural (excl. sponsor identity)** | 0.816 | 0.005 | 0.814 | 0.152 | 17,534 |
| **Baseline (Phase only)** | 0.775 | – | 0.782 | – | 17,534 |

**Table S8. Estimated Sunk Research Investment Associated With Silent Trials by Clinical Phase**.

| Trial Phase | Silent Trials (N) | Estimated Cost (USD) | Cost (Billions USD) |
|---|---|---|---|
| Unknown | 200 | $4,318,500,000 | 4.32 |
| Phase 3 | 302 | $4,139,370,000 | 4.14 |
| Phase 1 | 3,762 | $3,258,800,000 | 3.26 |
| Phase 2 | 459 | $2,509,955,000 | 2.51 |
| Phase 4 | 201 | $667,470,000 | 0.67 |
| Phase 1/2 | 60 | $179,520,000 | 0.18 |
| Phase 2/3 | 15 | $176,360,000 | 0.18 |
| Early Phase 1 | 18 | $12,520,000 | 0.01 |
| Unclassified | 2 | -- | -- |
| Total | 5,019 | $15,261,995,000 | 15.26 |

**Two trials with incomplete phase classification were included in the "Unclassified" category to ensure consistency with the overall silent trial count reported in the main manuscript.**

**Table S9. Estimated Economic Burden of Silent Trials by Sponsor**

| Sponsor | Estimated investment, billions USD |
|---|---|
| **Roche/Genentech** | 3.525 |
| **Sanofi** | 2.078 |
| **Novartis** | 1.776 |
| **Amgen** | 1.111 |
| **Bayer** | 1.086 |
| **Janssen (Johnson & Johnson)** | 0.902 |
| **Pfizer** | 0.858 |
| **AstraZeneca** | 0.566 |
| **GSK** | 0.530 |
| **Takeda** | 0.519 |
| **Bristol-Myers Squibb** | 0.444 |

| | |
|---|---|
| **AbbVie** | 0.409 |
| **Merck & Co.** | 0.368 |
| **Novo Nordisk** | 0.324 |
| **Boehringer Ingelheim** | 0.240 |
| **Eli Lilly** | 0.176 |
| **Teva** | 0.162 |
| **Gilead** | 0.121 |
| **Viatris/Mylan** | 0.046 |
| **Merck KGaA** | 0.021 |

**Estimated investment represents the participant-level sunk research investment associated with silent trials within each sponsor portfolio. Values are ranked in descending order of estimated burden.**

**Table S10. Dissemination Rates by Trial Phase and Sponsor**.

| Sponsor | Early Phase 1, % | Phase 1, % | Phase 1/2, % | Phase 2, % | Phase 2/3, % | Phase 3, % | Phase 4, % | Unknown phase, % |
|---|---|---|---|---|---|---|---|---|
| AbbVie | — | 46.8 | 90.0 | 87.7 | 100.0 | 93.8 | 91.3 | — |
| Amgen | 0.0 | 55.6 | 70.0 | 91.6 | 100.0 | 100.0 | 97.3 | 50.0 |
| AstraZeneca | 25.0 | 40.5 | 100.0 | 93.9 | 100.0 | 99.3 | 98.9 | 75.0 |
| Bayer | — | 22.5 | 60.0 | 65.3 | 80.0 | 79.8 | 70.4 | 2.8 |
| Bristol-Myers Squibb | — | 24.0 | 80.0 | 82.6 | 60.0 | 95.2 | 100.0 | 60.0 |
| Boehringer Ingelheim | 100.0 | 62.5 | 100.0 | 97.0 | 100.0 | 99.5 | 100.0 | 83.3 |
| Eli Lilly | 100.0 | 72.0 | 100.0 | 97.8 | 77.8 | 99.6 | 100.0 | 40.0 |

| | | | | | | | | |
|---|---|---|---|---|---|---|---|---|
| Gilead | — | 62.7 | 83.3 | 85.6 | 100.0 | 100.0 | 86.7 | — |
| GSK | — | 40.3 | 100.0 | 92.5 | 99.4 | 97.0 | 92.3 | — |
| Janssen (Johnson & Johnson) | 30.8 | 25.9 | 89.5 | 85.4 | 83.3 | 90.9 | 69.0 | 81.4 |
| Merck & Co. | 100.0 | 70.1 | 85.7 | 89.0 | 100.0 | 97.3 | 88.5 | 100.0 |
| Merck KGaA | 75.0 | 100.0 | 100.0 | 100.0 | 100.0 | 100.0 | 100.0 | — |
| Novartis | — | 30.5 | 63.8 | 82.3 | 84.0 | 92.3 | 84.4 | 80.0 |
| Novo Nordisk | 0.0 | 31.3 | 100.0 | 78.4 | 97.9 | 97.6 | 100.0 | — |
| Pfizer | 50.0 | 42.9 | 100.0 | 89.2 | 100.0 | 98.0 | 94.3 | 64.3 |
| Roche/Genentech | 100.0 | 42.5 | 100.0 | 80.5 | 33.3 | 90.0 | 87.0 | 54.2 |
| Sanofi | — | 33.3 | 62.2 | 71.6 | 66.7 | 70.2 | 65.9 | 33.3 |
| Takeda | — | 83.1 | 100.0 | 86.5 | 91.7 | 91.9 | 86.4 | 100.0 |
| Teva | — | 35.0 | 50.0 | 88.5 | 100.0 | 97.2 | 100.0 | — |
| Viatris/Mylan | — | 36.4 | 100.0 | 100.0 | — | 98.0 | 90.3 | 100.0 |

**Table S11. Sponsor-Level Operational Complexity Statistics and Silent Trial Counts**

| Sponsor | Trials, n | Mean OCI | Median OCI | SD of OCI | Dissemination rate, % | Silent trials, n |
|---|---|---|---|---|---|---|
| AbbVie | 493 | 0.140 | -0.157 | 0.981 | 80.1 | 98 |
| Amgen | 470 | 0.253 | -0.015 | 1.038 | 81.7 | 86 |
| AstraZeneca | 1,263 | -0.052 | -0.453 | 1.046 | 67.6 | 409 |
| Bayer | 763 | -0.091 | -0.479 | 1.061 | 44.3 | 425 |
| Bristol-Myers Squibb | 701 | 0.057 | -0.316 | 1.097 | 56.3 | 306 |
| Boehringer Ingelheim | 1,001 | -0.111 | -0.678 | 1.100 | 76.7 | 233 |
| Eli Lilly | 1,170 | -0.067 | -0.448 | 0.972 | 84.4 | 182 |

| | | | | | | |
|---|---|---|---|---|---|---|
| Gilead | 354 | 0.337 | 0.283 | 0.898 | 85.6 | 51 |
| GSK | 2,030 | -0.125 | -0.410 | 0.920 | 74.6 | 515 |
| Janssen (Johnson & Johnson) | 1,379 | -0.174 | -0.459 | 0.902 | 62.2 | 521 |
| Merck & Co. | 1,000 | -0.307 | -0.493 | 0.790 | 84.1 | 159 |
| Merck KGaA | 207 | -0.128 | -0.419 | 0.955 | 88.4 | 24 |
| Novartis | 1,627 | 0.348 | 0.192 | 0.945 | 76.8 | 377 |
| Novo Nordisk | 588 | 0.123 | -0.419 | 1.103 | 62.1 | 223 |
| Pfizer | 1,683 | -0.161 | -0.562 | 1.002 | 66.0 | 572 |
| Roche/Genentech | 1,005 | 0.301 | 0.065 | 1.064 | 71.0 | 291 |
| Sanofi | 1,053 | 0.091 | -0.149 | 0.949 | 60.6 | 415 |
| Takeda | 442 | -0.013 | -0.210 | 0.887 | 88.0 | 53 |
| Teva | 153 | 0.408 | 0.473 | 0.964 | 78.4 | 33 |
| Viatris/Mylan | 155 | -0.117 | -0.494 | 0.961 | 70.3 | 46 |

**Table S12. Manual Validation Metrics for Automated Dissemination Classification**

| OCI Tertile | N | Concordance | κ | FNR |
|---|---|---|---|---|
| Low | 115 | 91.3% | 0.82 | 11.2% |
| Medium | 115 | 92.2% | 0.84 | 9.8% |
| High | 114 | 92.9% | 0.86 | 9.1% |
| Total | 344 | 92.1% | 0.84 | 10.1% |

**Table S13. Sensitivity Analyses for the Association Between OCI and Trial Dissemination**

| Analysis Model | Adjusted OR | p-value |
|---|---|---|
| Phase 1 trials only | 2.40 | <0.001 |
| Phase 2 trials only | 1.57 | <0.001 |
| Phase 3 trials only | 2.94 | <0.001 |

| | | |
|---|---|---|
| **Follow-up ≥24 months** | **4.56** | **<0.001** |

Sensitivity analyses evaluating the robustness of the association between the Operational Complexity Index (OCI) and trial dissemination. Phase-stratified models were estimated to assess whether the OCI association persisted within individual clinical development phases. An additional sensitivity analysis restricted the cohort to trials with ≥24 months of follow-up after primary completion to account for legitimate reporting windows and publication lag.

**Table S14. Probabilistic Bias Analysis Adjusting for False-Negative Outcome Misclassification□**

| Analysis | Mean_OR | CI_low | CI_high |
|---|---|---|---|
| **PBA (10.1% False Negative)** | 4.1854 | 4.071254 | 4.302186 |

**Table S15. Principal Component Analysis of OCI Components**

| Variable | PC1 Loading |
|---|---|
| **log_enrollment** | **0.54** |
| **log_sites** | **0.60** |
| **log_countries** | **0.59** |

**Variance explained by PC1: 82.4%; Cronbach's alpha: 0.83**

**Table S16. Variance Inflation Factors for OCI Components**

| OCI Component | VIF |
|---|---|
| **log_enrollment** | **1.88** |
| **log_sites** | **2.31** |
| **log_countries** | **2.24** |

**Table S17. Comparison of Single-Component and Composite OCI Prediction Models**

| Model | Cross-validated AUC (mean) | Holdout AUC | Brier score |
|---|---|---|---|
| **Phase-only model** | **0.776** | — | — |
| **Enrollment-only model** | **0.757** | — | — |

| | | | |
|---|---|---|---|
| **Site-count-only model** | **—** | **0.796** | **—** |
| **Country-count-only model** | **—** | **0.796** | **—** |
| **Enrollment-component-only model** | **—** | **0.796** | **—** |
| **OCI structural model** | **0.816** | **0.814** | **0.152** |
| **Full model (OCI + sponsor identity)** | **0.858** | **0.857** | **0.134** |

**Table S18.Threshold-Based Classification Performance of the Predictive Model**

| Threshold | Sensitivity | Specificity | PPV | NPV |
|---|---|---|---|---|
| **0.30** | 0.798 | 0.748 | 0.559 | 0.902 |

*PPV = positive predictive value; NPV = negative predictive value. Thresholds represent predicted probabilities used to classify trials as being at risk of non-dissemination.*

**Table S19. Sponsor-Specific Associations Between OCI and Trial Dissemination**

| Sponsor | Trials, n | Odds ratio | 95% CI | p-value | Significance |
|---|---|---|---|---|---|
| **Teva** | **153** | **0.449** | **—** | **—** | **ns** |
| **Takeda** | **442** | **0.894** | **—** | **—** | **ns** |
| **Merck KGaA** | **207** | **1.021** | **—** | **—** | **ns** |
| **Boehringer Ingelheim** | **1,001** | **1.056** | **—** | **—** | **ns** |
| **Amgen** | **470** | **1.281** | **0.828–1.983** | **0.267** | **ns** |
| **Gilead** | **354** | **1.308** | **—** | **—** | **ns** |
| **Eli Lilly** | **1,170** | **1.361** | **0.988–1.874** | **0.059** | **ns** |

| | | | | | |
|---|---|---|---|---|---|
| **Pfizer** | **1,819** | **1.400** | **1.155–1.695** | **<0.001** | ******* |
| **Novartis** | **1,627** | **1.417** | **—** | **—** | **ns** |
| **AstraZeneca** | **1,263** | **1.502** | **1.164–1.937** | **0.002** | ****** |
| **Novo Nordisk** | **588** | **1.596** | **1.081–2.357** | **0.019** | ***** |
| **AbbVie** | **493** | **1.674** | **1.152–2.434** | **0.007** | ****** |
| **Janssen (Johnson & Johnson)** | **1,379** | **1.677** | **1.361–2.067** | **<0.001** | ******* |
| **Roche/Genentech** | **1,005** | **1.805** | **—** | **—** | **ns** |
| **Sanofi** | **1,053** | **1.869** | **1.583–2.208** | **<0.001** | ******* |
| **GSK** | **2,030** | **2.197** | **—** | **—** | **ns** |
| **Bayer** | **763** | **2.241** | **—** | **—** | **ns** |
| **Merck & Co.** | **996** | **2.308** | **—** | **—** | **ns** |
| **Bristol-Myers Squibb** | **701** | **2.716** | **2.020–3.651** | **<0.001** | ******* |

**Table S20. Schoenfeld Residual Tests for the Proportional Hazards Assumption**

| Covariate | Test statistic | p-value | Penalizer used |
|---|---|---|---|
| **OCI** | **58.046** | **<0.001** | **0.1** |
| **Post-FDAAA era** | **96.524** | **<0.001** | **0.1** |

p-values are based on Schoenfeld residual tests of the proportional hazards assumption. A non-zero test statistic with small p-value indicates evidence against the proportional hazards assumption.

**Table S21. Bootstrap Holdout AUC for the Predictive Model**

| Model | Holdout AUC | Bootstrap 95% CI | Test set, n | Bootstrap resamples, n |
|---|---|---|---|---|
| **Full model (OCI, sponsor, and covariates)** | 0.796 | 0.782–0.809 | 5,262 | 1,000 |

AUC = area under the receiver operating characteristic curve. Confidence intervals were estimated using bootstrap resampling of the holdout set.

**Table S22. Sensitivity Analysis of Research Investment Using Actual Versus Planned Enrollment**

| Enrollment source | Trials, n | Total cost, USD | Total cost, billions USD |
|---|---|---|---|
| **Actual enrollment** | **4,810** | **14.9 billion** | **14.9** |
| **Planned enrollment** | **209** | **303 million** | **0.3** |
| **All trials** | **5,019** | **15.2 billion** | **15.2** |

**Table S23.Registry Results Posting Rates by OCI Tertile**

| OCI Tertile | N | Registry Posted, n (%) | Silent, n (%) | aOR | 95% CI | p-value |
|---|---|---|---|---|---|---|
| **Low** | **5,848** | **1,909 (32.6%)** | **3,939 (67.4%)** | **Reference** | **—** | **—** |
| **Medium** | **5,843** | **3,721 (63.7%)** | **2,122 (36.3%)** | **—** | **—** | **—** |
| **High** | **5,846** | **5,003 (85.6%)** | **843 (14.4%)** | **3.46** | **3.30–3.63** | **<0.001** |

aOR = adjusted odds ratio from binary logistic regression with OCI (standardized) and post-FDAAA status as covariates. Registry results posting is mandated under the FDA Amendments Act (FDAAA 801) irrespective of trial outcome. Silent = no registry posting and no PubMed-indexed publication identified.

**Table S24. Table S25. Sensitivity Analyses of Estimated Participant-Level Research Investment Associated With Silent Trials**

| Scenario | Silent trials, n | Assumption for unknown phase (n = 200) | Estimated investment, USD |
|---|---|---|---|
| **Primary estimate** | **5,019** | **Phase 1 benchmark ($20,000 per participant)** | **$15.26 billion** |
| **Sensitivity 1: Enrollment capped at the 99th percentile (777 participants)** | **5,019** | **Enrollment capped at 777 participants** | **$11.41 billion** |
| **Sensitivity 2: Median enrollment imputed (36 participants)** | **5,019** | **Median of known-phase trials (36 participants)** | **$11.12 billion** |
| **Sensitivity 3: Unknown phase excluded** | **4,819** | **Excluded entirely** | **$10.94 billion** |

Estimated investment represents participant-level sunk research investment associated with silent trials under alternative assumptions for studies with unknown phase or extreme enrollment values.

**Table S25. Table S26. Estimated Participant-Level Research Investment Associated With Silent Trials by Clinical Phase**

| Clinical phase | Silent trials, n | Estimated investment, USD |
|---|---|---|
| Early Phase 1 | 18 | $0.01 billion |
| Phase 1 | 3,762 | $3.26 billion |
| Phase 1/2 | 60 | $0.18 billion |
| Phase 2 | 459 | $2.51 billion |
| Phase 2/3 | 15 | $0.18 billion |
| Phase 3 | 302 | $4.14 billion |
| Phase 4 | 201 | $0.67 billion |
| Unknown phase | 200 | $4.32 billion |
| Total | 5,019 | $15.26 billion |

**Totals reflect the primary economic-burden estimate.**

**Table S26. Table S26. Cost Benchmarks and Assumptions Used to Estimate Participant-Level Research Investment**

| Clinical phase | Cost benchmark per participant, USD | Assumption |
|---|---|---|
| Early Phase 1 | $20,000 | Benchmark applied to early human-phase studies |
| Phase 1 | $20,000 | Benchmark applied to phase 1 studies |
| Phase 1/2 | $27,500 | Intermediate benchmark |
| Phase 2 | $35,000 | Benchmark applied to phase 2 studies |
| Phase 2/3 | $40,000 | Intermediate benchmark |
| Phase 3 | $45,000 | Benchmark applied to phase 3 studies |
| Phase 4 | $15,000 | Lower benchmark applied to post-marketing studies |
| Unknown phase | $25,000 | Conservative fallback assumption |

**Table S27. Sponsor-Specific OCI Effects on Trial Dissemination**

| Sponsor | N | aOR | 95% CI | p-value |
|---|---|---|---|---|
| GSK | 2,030 | 24.05 | 16.49–35.07 | <0.001 |
| Merck Co. | 1,000 | 15.69 | 8.54–28.84 | <0.001 |
| ViatrisMyllan | 155 | 11.66 | 3.79–35.85 | <0.001 |
| AstraZeneca | 1,263 | 11.13 | 7.93–15.62 | <0.001 |
| Boehringer Ingelheim | 1,001 | 9.34 | 5.80–15.03 | <0.001 |
| Pfizer | 1,683 | 7.43 | 5.79–9.54 | <0.001 |
| Novo Nordisk | 588 | 7.59 | 5.18–11.11 | <0.001 |
| Bristol-Myers Squibb | 701 | 6.83 | 5.05–9.24 | <0.001 |
| Merck KGaA | 207 | 6.10 | 1.89–19.63 | 0.002 |
| Eli Lilly | 1,170 | 5.84 | 3.95–8.65 | <0.001 |
| Janssen (J&J) | 1,379 | 5.26 | 4.15–6.65 | <0.001 |
| Teva | 153 | 5.31 | 2.75–10.26 | <0.001 |
| Amgen | 470 | 5.05 | 3.21–7.95 | <0.001 |
| Bayer | 763 | 4.93 | 3.88–6.27 | <0.001 |
| Sanofi | 1,053 | 3.69 | 3.01–4.52 | <0.001 |
| RocheGenentech | 1,005 | 2.77 | 2.30–3.33 | <0.001 |
| AbbVie | 493 | 2.75 | 1.93–3.91 | <0.001 |
| Novartis | 1,627 | 3.05 | 2.55–3.65 | <0.001 |
| Gilead | 354 | 3.17 | 2.01–5.00 | <0.001 |
| Takeda | 442 | 1.70 | 1.13–2.57 | 0.011 |


1. Donner, A. and M. Eliasziw, A goodness-of-fit approach to inference procedures for the kappa statistic: Confidence interval construction, significance-testing and sample size estimation. Statistics in medicine, 1992. **11**(11): p. 1511-1519.
